# Comparison of the Expert Guidelines With Artificial Intelligence-Driven Echocardiographic Assessment of Diastolic Function

**DOI:** 10.64898/2026.04.23.26350072

**Authors:** Márton Tokodi, Nobuyuki Kagiyama, Ambarish Pandey, Yutaka Nakamura, Yuka Akama, Sachiko Takamatsu, Misako Toki, Takeshi Kitai, Taiji Okada, Carolyn S. P. Lam, Naveena Yanamala, Partho P. Sengupta

## Abstract

**Backgound:** Accurate assessment of diastolic function and left ventricular (LV) filling pressure is central to heart failure diagnosis and risk stratification. Contemporary guideline algorithms rely on complex parameters that are not consistently available in routine clinical practice.

**Objective:** To compare the diagnostic and prognostic performance of the 2016 American Society of Echocardiography/European Association of Cardiovascular Imaging (ASE/EACVI) and 2025 ASE guidelines with a deep learning model based on routinely acquired echocardiographic variables.

**Methods:** This study evaluated the guideline-based algorithms and a deep learning model in participants from the Atherosclerosis Risk in Communities (ARIC) cohort (n=5450) for prognostication and two invasive hemodynamic validation cohorts from the United States (n=83) and Japan (n=130) for detection of elevated left ventricular filling pressure.

**Results:** In the ARIC cohort, the deep learning model demonstrated superior prognostic performance compared with the 2016 and 2025 guidelines (C-index: 0.676 vs. 0.638 and 0.602, respectively; both p<0.001). Similar findings were observed among participants with preserved ejection fraction (C-index: 0.660 vs. 0.628 and 0.590; both p<0.001), with improved performance compared with the H_2_FPEF score (C-index: 0.660 vs. 0.607; p<0.001). In the US hemodynamic validation cohort, the deep learning model showed higher diagnostic performance than the 2025 guidelines (AUC: 0.879 vs. 0.822; p=0.041) and similar performance compared with the 2016 guidelines (AUC: 0.879 vs. 0.812; p=0.138). In the Japanese hemodynamic validation cohort, the deep learning model outperformed both guidelines (AUC: 0.816 vs. 0.634 and 0.694; both p<0.05).

**Conclusions:** A deep learning model leveraging routinely available echocardiographic parameters demonstrated improved diagnostic and prognostic performance compared with contemporary guideline-based approaches, potentially offering a scalable alternative for assessing diastolic function and left ventricular filling pressures.

## INTRODUCTION

Left ventricular (LV) diastolic dysfunction (DD) is a common and prognostically significant abnormality in patients with cardiovascular diseases, contributing to symptoms of heart failure (HF), reduced exercise tolerance, and adverse outcomes, even in the presence of preserved LV ejection fraction (LVEF) (1,2). Because elevated LV filling pressure (LVFP) is the hemodynamic substrate underlying these outcomes, accurate assessment of diastolic function is central to contemporary HF care and risk stratification.

To guide this assessment, the American Society of Echocardiography (ASE) and the European Association of Cardiovascular Imaging (EACVI) developed guidelines that define stepwise decision algorithms based on echocardiographic parameters (3–6). The 2016 recommendations provided a pragmatic framework but showed modest correlation with invasively measured LVFP and a high rate of indeterminate results (7–11). The updated 2025 ASE guidelines introduced additional Doppler and strain parameters, along with refined decision pathways, to reduce indeterminate classifications and improve diagnostic accuracy (5,12). However, implementation depends on complex measures that are not consistently available in routine clinical practice (12,13).

Rule-based guideline algorithms rely on fixed cutoffs and may not capture the multivariable, nonlinear, and age-dependent nature of diastolic physiology (14). In contrast, machine learning and deep learning (DL) approaches can integrate multiple echocardiographic features, model age-related variation, and improve prediction of outcomes and physiologic states across diverse cohorts (15–18).

Accordingly, we aimed to compare the 2025 ASE and 2016 ASE/EACVI guidelines with a validated DL model in participants of the Atherosclerosis Risk in Communities (ARIC) cohort study for predicting HF hospitalization or all-cause death, and to evaluate their diagnostic performance for detecting elevated LVFP in two geographically distinct cohorts from the United States (US) and Japan. We hypothesized that the DL model, which was designed to extract latent physiologic traits from standard echocardiographic variables, would match or even surpass the diagnostic and prognostic performance of contemporary guideline-based algorithms across diverse populations, even in the presence of missing complex Doppler or strain measurements.

## METHODS

### Validation cohort for predicting clinical outcomes – the ARIC cohort

We validated the prognostic value of the guidelines and the DL model using data collected in the ARIC cohort study (19). We analyzed the data of those participants who underwent a transthoracic echocardiographic examination at visit 5 between 2011 and 2013 and whose data were available in the Biologic Specimen and Data Repository Information Coordinating Center (BioLINCC) database (Figure 1). From the 5,576 participants who fulfilled these criteria, we excluded those with any degree of mitral stenosis (n=33), severe mitral regurgitation (n=2), documented mitral annular calcification (n=2), a history of heart transplantation (n=1), a history of LV assist device implantation (n=8), previous mitral valve interventions (n=20), or a prosthetic valve at any position (n=47). In addition, we also excluded participants with more than three missing values in key echocardiographic variables (i.e., the input features of the DL model; n=31) or no follow-up data (n=3). The echocardiographic protocol of visit 5 has been published previously (20) and is described briefly in the Supplemental Methods. H_2_FPEF score was calculated for participants with preserved LVEF (≥50%) according to the original published algorithm (21), as detailed in the Supplemental Methods. The primary outcome of interest was the composite of HF hospitalization or all-cause death, but the main results are also reported for HF hospitalization and all-cause death separately. The time to event was measured from the date of the echocardiographic examination at visit 5.

**Figure 1.**
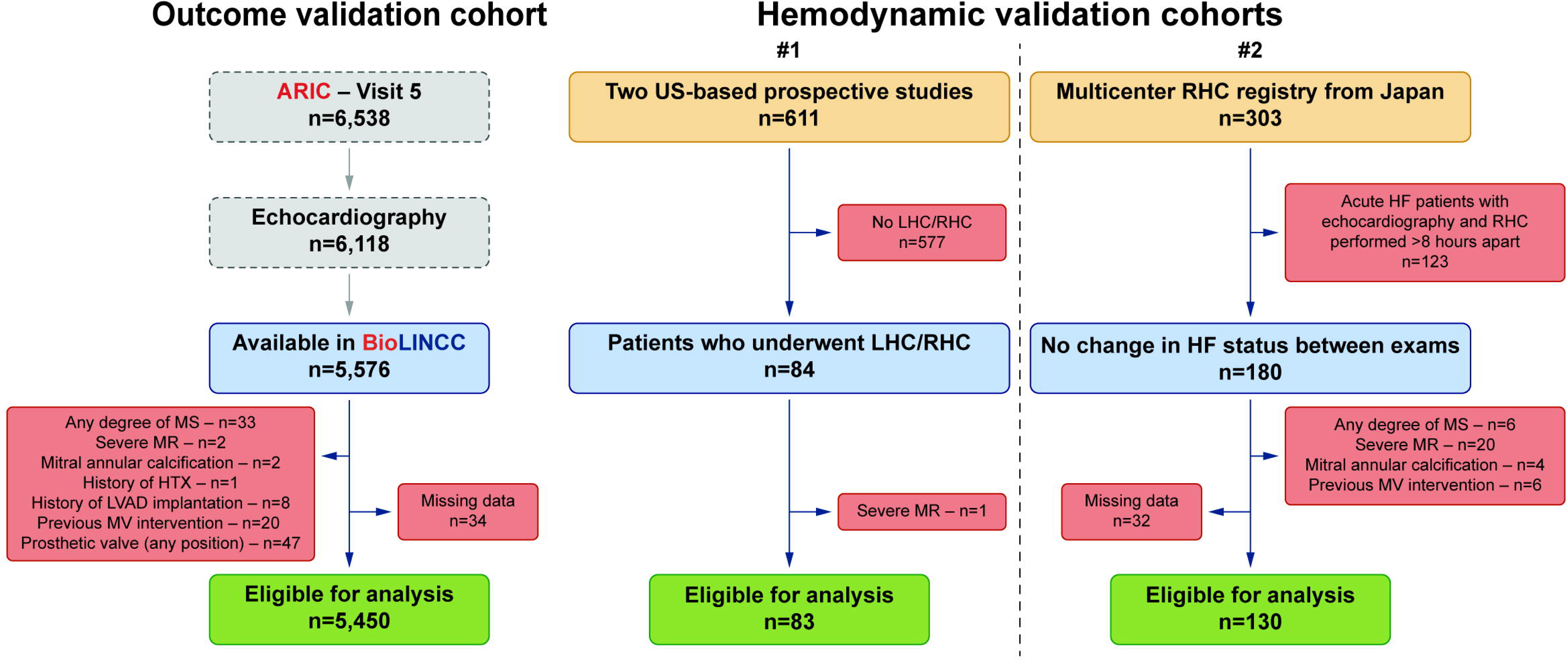
Patient selection flowcharts of the three cohorts included in the study ARIC – Atherosclerosis Risk In Communities, BioLINCC – Biologic Specimen and Data Repository Information Coordinating Center, HF – heart failure, HTX – heart transplantation, LHC – left heart catheterization, LVAD – left ventricular assist device, MR – mitral regurgitation, MS – mitral stenosis, MV – mitral valve, RHC – right heart catheterization

### Validation cohorts for detecting elevated LVFP

We validated the guidelines and the DL model against invasively measured LVFP in two distinct cohorts of patients with paired echocardiographic and invasive hemodynamic studies (Figure 1). The first cohort comprised patients with varying degrees of systolic and diastolic dysfunction who underwent transthoracic echocardiography and invasive hemodynamic assessment either with left heart catheterization (LHC) or right heart catheterization (RHC) when enrolled in two prospective studies at West Virginia University (Morgantown, West Virginia) (22,23). From the 84 participants who fulfilled these criteria, we excluded those with severe mitral regurgitation (n=1). None of the patients met the other exclusion criteria listed for the ARIC cohort, as they were prospectively enrolled in clinical studies with inclusion and exclusion criteria previously defined to permit echocardiographic assessment of diastolic function (22,23). The protocols of echocardiographic examinations and invasive hemodynamic evaluations have been described previously (22,23).

The second hemodynamic validation cohort was derived from a Japanese retrospective multicenter registry that included 303 adults (≥20 years) who underwent both transthoracic echocardiography and RHC within 24 hours at Juntendo University (Tokyo, Japan), the Sakakibara Heart Institute of Okayama (Okayama, Japan), or the Kobe City Medical Center General Hospital (Kobe, Japan). To minimize errors resulting from changes in HF status between the two examinations, patients hospitalized for acute HF in whom echocardiography and RHC were performed more than 8 hours apart were excluded from the present study (n=123). Patients with any degree of mitral stenosis (n=6), severe mitral regurgitation (n=20), mitral annular calcification (n=4), previous mitral valve interventions (n=6), more than three missing values in key echocardiographic variables (i.e., the input features of the DL model; n=28), or missing pulmonary capillary wedge pressure (n=4) were also excluded. Echocardiographic examinations were performed in accordance with the current ASE/EACVI guidelines (24). RHC was performed via the jugular or femoral vein using a 5–8 Fr balloon-tipped, fluid-filled Swan-Ganz catheter (Edwards Lifesciences, Irvine, California, USA). Intracardiac pressures, including pulmonary capillary wedge pressure, were assessed under fluoroscopic guidance after calibration with zero being positioned at the mid-thoracic line.

In the hemodynamic validation cohorts, elevated LVFP was defined as an RHC-derived pulmonary capillary wedge pressure or an LHC-derived LV pre-atrial contraction pressure greater than 15 mmHg.

### Guideline-based echocardiographic assessment of diastolic function

Diastolic function was assessed in all three cohorts according to the 2016 ASE/EACVI guidelines (4) and the 2025 ASE guidelines (5).

Using the 2016 ASE/EACVI guidelines, patients were classified into five categories: (1) normal diastolic function, (2) indeterminate diastolic function, (3) DD with normal LAP (i.e., grade 1 DD), (4) DD with indeterminate LAP, and (5) DD with elevated LAP (i.e., grade 2 or 3 DD).

According to the 2025 ASE guidelines, patients were assigned to one of four categories: (1) normal diastolic function, (2) DD with normal LAP, (3) DD with indeterminate LAP, and (4) DD with elevated LAP. Although the 2025 ASE guidelines were developed to potentially reduce the number of cases with indeterminate assessment, patients were still classified as having DD with indeterminate LAP (1) if they were in sinus rhythm, only early diastolic mitral annular velocity (e’) was reduced among the primary parameters, and the ratio of the early and late mitral inflow velocities (E/A) was unavailable, (2) if they were in sinus rhythm and additional parameters were required to assess LAP but none were available, (3) if they had non-cardiac pulmonary hypertension and E/A was unavailable, (4) if they had non-cardiac pulmonary hypertension, LAP could not be assessed solely based on E/A, left atrial reservoir strain (LARS) was unavailable, and lateral E/e’ was 8–13, (5) if they were in atrial fibrillation and less than three of the primary parameters were available, and (6) if they were in atrial fibrillation and exactly two of the primary criteria were met, but less than two of the secondary parameters were available.

The availability of the echocardiographic parameters used for diastolic function assessment is reported in Supplemental Table 1.

### DL-based assessment of diastolic function

We used our previously published and extensively validated DL model that assesses diastolic function based on nine routinely measured echocardiographic parameters: LVEF, LV mass index, left atrial volume index, E, A, E/A, septal e’, septal E/e’, and tricuspid regurgitation peak velocity (15,25). The output of the DL model is a single numeric value for each subject, denoting the probability of DD. One of the algorithm’s significant advantages is its ability to provide accurate diagnostic and prognostic predictions even when input echocardiographic variables contain missing values. Further details on the DL model and its external validation are provided in the Supplemental Methods and have been published previously (15). The model is publicly available online (26).

### Statistical analysis

Continuous variables are expressed as median (interquartile range), while categorical variables are reported as frequencies and percentages. The normality of continuous variables was checked using the Shapiro-Wilk test. The characteristics of patient subgroups were compared using unpaired Student’s t-test or Mann-Whitney U test for continuous variables and Chi-squared or Fisher’s exact test for categorical variables, as appropriate. The event-free survival of the subgroups was visualized using Kaplan-Meier curves, and pairwise log-rank tests with the Benjamini-Hochberg adjustment were applied for comparison. For the composite endpoint and all-cause death, Cox proportional hazards models were used to compute hazard ratios (HRs) with 95% confidence intervals (CIs). For HF hospitalization, subdistribution HRs with 95% CIs were estimated using Fine-Gray competing risks regression, with all-cause death treated as the competing event. To compare the prognostic value of the DL model and the guidelines, Harrell’s C-indices of Cox regression models in the same patients were compared using the *CompareC* R package (27). Bootstrap resampling was applied to calculate P-values when comparing C-indices derived from Cox regression models in different patient subsets or from Fine-Gray competing risk models. Time-dependent receiver operating characteristic (ROC) analysis was also performed to support and strengthen the results of the C-index comparisons. The performance of the guidelines and the DL model in detecting elevated LVFP was assessed using the area under the ROC curve (AUC). Because the guideline-based classifications were nominal, univariable logistic regression models were fitted in which the classifications were used as predictors of the binary reference standard (i.e., elevated LVFP). Through this approach, the nominal classifications were converted into continuous probability estimates, enabling the calculation of AUCs and allowing for direct comparison with the DL model using DeLong tests. A P-value of <0.05 was considered statistically significant. All statistical analyses were performed in R (version 4.5.2, R Foundation for Statistical Computing, Vienna, Austria).

### Ethical approval

All participants of the ARIC cohort study provided written informed consent, and the institutional review boards associated with each field center approved the study protocol. The two US-based prospective studies used for hemodynamic validation were approved by the corresponding institutional review boards, and written informed consent was obtained from all patients. Data collection and analysis for the multicenter Japanese registry were approved by the institutional review boards of each center. Given the retrospective observational design, written informed consent was waived, and an opt-out consent procedure was implemented. The protocol for the current analysis adheres to the principles outlined in the Declaration of Helsinki and was approved by the Institutional Review Board of Rutgers Biomedical and Health Sciences (study identification number: Pro2021001505). Methods and results are reported in compliance with the updated Proposed Requirements for Cardiovascular Imaging-Related Multimodal-AI Evaluation (PRIME 2.0) checklist (Supplemental Table 2) (28).

## RESULTS

### Prognostic value in the entire ARIC cohort

Of the 5,450 ARIC study participants eligible for analysis (75 [71–79] years, 43% male), 1,320 (24%) reached the composite endpoint of HF hospitalization or all-cause death over 6.5 (6.1–7.0) years. As shown in Figure 2, reclassifications were observed between the three approaches, with significantly fewer patients classified as indeterminate using the 2025 guidelines than the 2016 guidelines (124 [2%] vs. 1,082 [20%], p<0.001). The clinical and echocardiographic characteristics of the ARIC cohort are reported in Table 1 and Supplemental Table 3.

**Figure 2.**
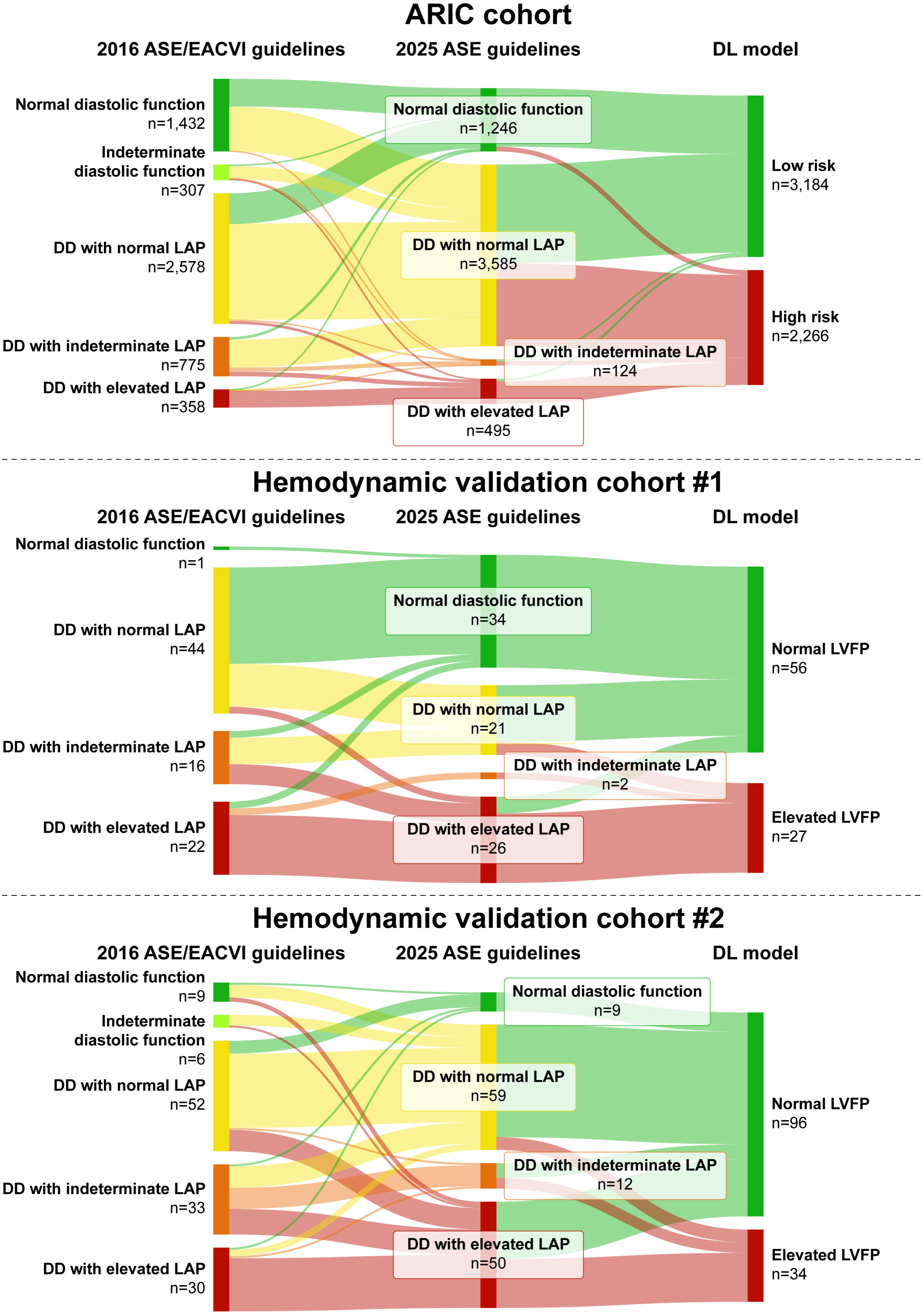
Sankey diagrams depicting the reclassification of patients between the guidelines and the DL model Given that the DL model was primarily trained for prognostication, a cutoff optimized for identifying elevated LVFP using Youden’s J statistic was applied instead of the default 0.5 threshold to dichotomize the predicted probabilities in the hemodynamic validation cohorts. Because the optimal cutoff depends on cohort characteristics, separate cutoffs were determined for the two hemodynamic validation cohorts. In the ARIC cohort, the default 0.5 threshold was used to classify patients as high risk or low risk. ASE – American Society of Echocardiography, DD – diastolic dysfunction, DL – deep learning, EACVI – European Association of Cardiovascular Imaging, LAP – left atrial pressure, LV – left ventricular, LVFP – left ventricular filling pressure; other abbreviations as in Figure 1.

**Table 1.** Clinical and echocardiographic characteristics of the three cohorts.

|  | ARIC cohort<br>n=5,450 | Hemodynamic<br>validation cohort #1<br>n=83 | Hemodynamic<br>validation cohort #2<br>n=130 |
| --- | --- | --- | --- |
| **Demographics, vitals, risk factors** |  |  |  |
| Age, years | 75 (71–79) | 61 (54–66) | 66 (51–75) |
| Male sex | 2,326 (43) | 56 (67) | 70 (54) |
| Black race | 1,049 (19) | 3 (4) | 0 (0) |
| BMI, kg/m<sup>2</sup> | 27.9 (24.9–31.5) | 30.8 (27.6–34.7) | 22.0 (19.4–24.7) |
| BSA, m<sup>2</sup> | 1.86 (1.70–2.01) | 2.11 (1.94–2.28) | 1.58 (1.43–1.73) |
| SBP, mmHg | 129 (118–141) | 133 (117–147) | 112 (100–126) |
| DBP, mmHg | 66 (59–74) | 77 (69–89) | 65 (57–76) |
| HR, 1/min | 64 (57–71) | 73 (65–85) | 70 (62–80) |
| Hypertension | 4,391 (81) | 82 (99) | 49 (38) |
| Diabetes | 2,004 (37) | 42 (51) | 30 (23) |
| Atrial fibrillation | 166 (3) | 6 (7) | 21 (16) |
| Chronic kidney disease | 1,500 (28) | 14 (17) | - |
| History of HF | 908 (17) | 30 (36) | - |
| History of CHD | 1,048 (19) | 67 (81) | 29 (22) |
| **Laboratory results** |  |  |  |
| BNP, pg/mL | - | 1,068 (425–2,170) | 168 (45–318) |
| NT-proBNP, pg/mL | 134 (69–264) | - | 326 (192–1,282) |
| **Echocardiographic parameters** |  |  |  |
| IVSd, mm | 10 (9–11) | 11 (11–14) | 10 (9–11) |
| LVPWd, mm | 9 (8–10) | 10 (9–12) | 10 (9–11) |
| LVIDd, mm | 44 (41–47) | 46 (42–50) | 47 (41–56) |
| LVIDs, mm | 26 (23–29) | 33 (29–35) | 31 (26–45) |
| LVMi, g/m<sup>2</sup> | 76.4 (66.1–89.1) | 99.0 (80.8–118.3) | 102.5 (81.5–141.8) |
| LVEDVi, mL/m<sup>2</sup> | 42.4 (36.4–49.7) | 57.3 (47.7–76.0) | 74.2 (56.2–97.5) |
| LVESVi, mL/m<sup>2</sup> | 14.4 (11.6–17.9) | 26.9 (19.6–40.3) | 32.3 (16.3–71.7) |
| LVEF, % | 65.8 (61.9–69.2) | 54.0 (38.0–59.5) | 61.9 (37.3–69.9) |
| LAVi, mL/m<sup>2</sup> | 24.7 (20.3–30.2) | 27.9 (20.7–38.7) | 35.5 (25.1–48.6) |
| LARS, % | - | 21.4 (11.0–30.5) | 17.2 (10.7–27.8) |
| Moderate or greater AS | 40 (1) | 3 (4) | 8 (9) |
| Moderate or greater AR | 22 (0) | 1 (1) | 10 (8) |
| Moderate MR | 98 (2) | 11 (13) | 21 (16) |
| E, cm/s | 65 (54–78) | 81 (63–95) | 70 (51–90) |
| E-DT, ms | 200 (173–230) | 202 (192–232) | 192 (150–238) |
| A, cm/s | 79 (67–91) | 70 (56–85) | 70 (49–84) |
| E/A | 0.8 (0.7–1.0) | 1.0 (0.8–1.5) | 0.9 (0.6–1.4) |
| IVRT, ms | - | 77 (70–90) | 89 (73–114) |
| e' (septal), cm/s | 5.5 (4.7–6.5) | 6.0 (4.7–7.3) | 5.5 (3.7–7.3) |
| E/e' (septal) | 11.6 (9.5–14.4) | 12.7 (9.5–20.0) | 13.1 (9.7–16.8) |
| e' (lateral), cm/s | 6.8 (5.6–8.2) | 8.0 (6.0–9.9) | 8.4 (6.7–11.4) |
| E/e' (lateral) | 9.5 (7.6–12.0) | 9.1 (6.9–14.6) | 8.3 (5.9–10.8) |
| e' (average), cm/s | 6.2 (5.3–7.3) | 7.5 (5.5–8.4) | 7.3 (5.6–9.2) |
| E/e' (average) | 10.6 (8.7–13.1) | 10.1 (8.0–17.8) | 9.2 (7.2–11.9) |
| TRV, m/s | 2.36 (2.18–2.55) | 2.54 (1.87–2.95) | 2.69 (2.24–3.20) |
| PASP, mmHg | - | 24 (13–37) | 37 (26–49) |
| 2016 ASE/EACVI guidelines |  |  |  |
| Normal diastolic function | 1,432 (26) | 1 (1) | 9 (7) |
| Indeterminate diastolic function | 307 (6) | 0 (0) | 6 (5) |
| DD with normal LAP | 2,578 (47) | 44 (53) | 52 (40) |
| DD with indeterminate LAP | 775 (14) | 16 (19) | 33 (25) |
| DD with elevated LAP | 358 (7) | 22 (27) | 30 (23) |
| 2025 ASE guidelines |  |  |  |
| Normal diastolic function | 1,246 (23) | 34 (41) | 9 (7) |
| DD with normal LAP | 3,585 (66) | 21 (25) | 59 (45) |
| DD with indeterminate LAP | 124 (2) | 2 (2) | 12 (9) |
| DD with elevated LAP | 495 (9) | 26 (31) | 50 (38) |
| DL-predicted probability of DD | 0.30 (0.04–0.86) | 0.95 (0.21–1.00) | 0.98 (0.60–0.99) |
| **Invasive hemodynamic assessment** |  |  |  |
| LVFP, mmHg | - | 11.1 (7.7–17.5) | 10.0 (7.0–14.8) |
| LVFP >15 mmHg | - | 25 (30) | 30 (23) |

Continuous variables are presented as median (interquartile range) and categorical variables as n (%).
A – late mitral inflow velocity, AR – aortic regurgitation, ARIC – Atherosclerosis Risk in Communities study, AS – aortic stenosis, ASE – American Society of Echocardiography, BMI – body mass index, BNP – brain natriuretic peptide, BSA – body surface area, CHD – coronary heart disease, DBP – diastolic blood pressure, DD – diastolic dysfunction, DL – deep learning, E – early mitral inflow velocity, EACVI – European Association of Cardiovascular Imaging, E-DT – E-wave deceleration time, e' – early diastolic mitral annular velocity, HF – heart failure, HR – heart rate, IVRT – isovolumic relaxation time, IVSd – thickness of the interventricular septum at end-diastole, LAP – left atrial pressure, LARS – left atrial reservoir strain, LAVi – left atrial volume index, LV – left ventricular, LVEDVi – left ventricular end-diastolic volume index, LVEF – left ventricular ejection fraction, LVESVi – left ventricular end-systolic volume index, LVFP – left ventricular filling pressure, LVIDd – left ventricular internal diameter at end-diastole, LVIDs – left ventricular internal diameter at end-systole, LVMI – left ventricular mass index, LVPWd – thickness of the left ventricular posterior wall at end-diastole, MR – mitral regurgitation, NT-proBNP – N-terminal pro-brain natriuretic peptide, PASP – pulmonary artery systolic pressure estimated by echocardiography, SBP – systolic blood pressure, TRV – tricuspid regurgitation peak velocity

The guideline-derived diastolic function categories, as well as the DL-predicted probabilities, were associated with the composite endpoint of HF hospitalization or all-cause death both in univariable and multivariable Cox regression analyses (Table 2 and Supplemental Tables 4 and 5). Moreover, both guideline-based classifications and the DL-predicted probability exhibited incremental prognostic value over N-terminal pro-brain natriuretic peptide (NT-proBNP) (Supplemental Table 6). Notably, the DL model demonstrated superior prognostic power over both guidelines in univariable Cox regression, with a C-index of 0.676 (95% CI: 0.660–0.692) vs. 0.638 (95% CI: 0.624–0.652) for the 2016 guidelines and 0.602 (95% CI: 0.588–0.616) for the 2025 guidelines (p<0.001 for both comparisons; Figure 3), even after adjustment for cardiovascular risk factors in multivariable Cox regression, with a C-index of 0.739 (95% CI: 0.725–0.753) vs. 0.732 (95% CI: 0.718–0.746) for the 2016 guidelines and 0.728 (95% CI: 0.714–0.742) for the 2025 guidelines (p=0.030 and p<0.001, respectively; Supplemental Figure 1). The 2016 guidelines achieved a higher C-index than the 2025 guidelines in univariable analysis (p<0.001; Figure 3) but not after adjustment for risk factors in multivariable analysis (p=0.058; Supplemental Figure 1). These findings were further supported by the results of the time-dependent ROC analyses (Supplemental Figure 2 and Supplemental Table 7). We also found consistent results across patient subgroups stratified by race, the presence of atrial fibrillation, history of heart failure or coronary heart disease, and elevated NT-proBNP levels (Supplemental Table 8). After excluding patients with indeterminate diastolic function or DD with indeterminate LAP, the DL model maintained superior prognostic discrimination compared to both guidelines, with a C-index of 0.676 (95% CI: 0.660–0.692) vs. 0.624 (95% CI: 0.609–0.639) for the 2016 guidelines and 0.590 (95% CI: 0.576–0.604) for the 2025 guidelines (p<0.001 for both comparisons), and the 2016 guidelines also outperformed the 2025 guidelines (p=0.002; Figure 3).

**Figure 3.**
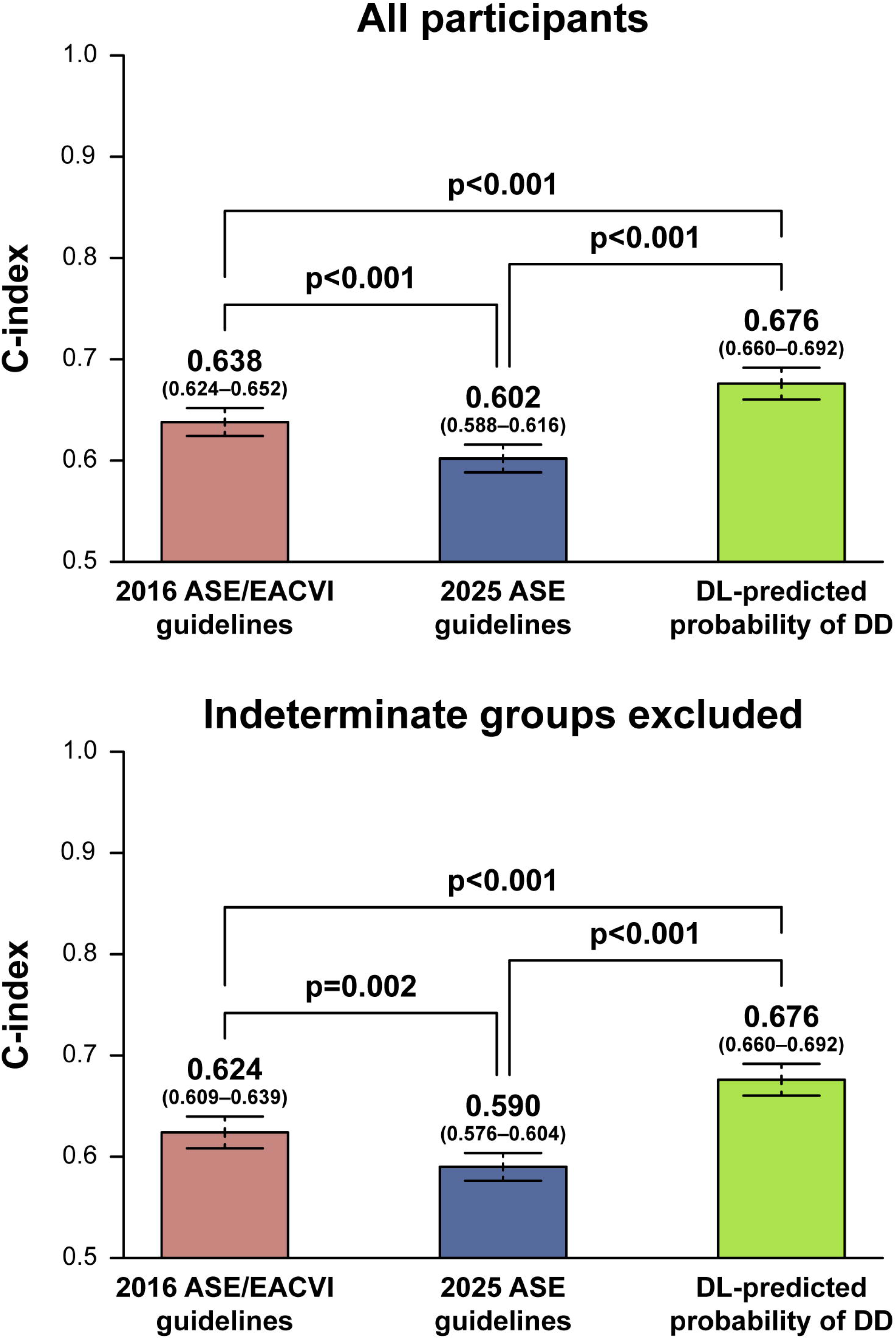
Prognostic value of the guidelines and the DL model for predicting the composite endpoint C-indices were calculated from univariable Cox regression models containing either the 2016 or 2025 guideline-based classification or the DL-predicted probability of DD. Models in the upper panel were evaluated in the entire ARIC cohort, whereas those in the lower panel were evaluated after excluding indeterminate groups. The two guidelines classified different patients as indeterminate; therefore, their performance was evaluated in overlapping but non-identical subsets after excluding indeterminate cases: the 2016 ASE/EACVI guidelines were evaluated in 67 patients, whereas the 2025 ASE guidelines were evaluated in 79 patients. P-values for C-index comparison were calculated using the *CompareC* R package (27) in the upper panel, whereas they were calculated using bootstrap resampling in the lower panel. C-indices are reported with 95% CIs. Abbreviation as in Figures 1 and 2.

**Table 2.** Univariable and multivariable Cox regression models predicting the composite endpoint of HF hospitalization or all-cause death in the ARIC cohort.

|  | Univariable models |  | Multivariable model #1<br>C-index: 0.732 (0.718–0.746) |  | Multivariable model #2<br>C-index: 0.728 (0.714–0.742) |  | Multivariable model #3<br>C-index: 0.739 (0.725–0.753) |  |
| --- | --- | --- | --- | --- | --- | --- | --- | --- |
|  | HR (95% CI) | P-value | HR (95% CI) | P-value | HR (95% CI) | P-value | HR (95% CI) | P-value |
| Age | 1.106 (1.095–1.117) | <0.001 | 1.080 (1.069–1.092) | <0.001 | 1.081 (1.069–1.092) | <0.001 | 1.075 (1.064–1.087) | <0.001 |
| Male sex | 1.505 (1.351–1.676) | <0.001 | 1.364 (1.219–1.526) | <0.001 | 1.342 (1.200–1.500) | <0.001 | 1.286 (1.150–1.438) | <0.001 |
| Hypertension | 2.006 (1.696–2.372) | <0.001 | 1.125 (0.945–1.341) | 0.186 | 1.161 (0.975–1.382) | 0.094 | 1.078 (0.905–1.285) | 0.400 |
| Diabetes | 1.710 (1.534–1.905) | <0.001 | 1.411 (1.263–1.577) | <0.001 | 1.417 (1.268–1.584) | <0.001 | 1.375 (1.230–1.537) | <0.001 |
| Atrial fibrillation | 2.769 (2.220–3.454) | <0.001 | 1.330 (1.032–1.715) | 0.028 | 1.475 (1.152–1.889) | 0.002 | 1.554 (1.240–1.949) | <0.001 |
| Chronic kidney disease | 1.902 (1.703–2.124) | <0.001 | 1.385 (1.235–1.552) | <0.001 | 1.404 (1.252–1.573) | <0.001 | 1.404 (1.253–1.574) | <0.001 |
| History of HF | 3.267 (2.912–3.666) | <0.001 | 2.019 (1.773–2.300) | <0.001 | 2.129 (1.872–2.422) | <0.001 | 2.037 (1.791–2.317) | <0.001 |
| History of CHD | 2.340 (2.085–2.627) | <0.001 | 1.209 (1.059–1.380) | 0.005 | 1.284 (1.127–1.463) | <0.001 | 1.206 (1.058–1.376) | 0.005 |
| History of non-cardiac PH | 3.761 (2.970–4.763) | <0.001 | 1.778 (1.393–2.270) | <0.001 | 1.427 (1.093–1.863) | 0.009 | 1.690 (1.324–2.157) | <0.001 |
| History of stroke | 2.203 (1.780–2.726) | <0.001 | 1.142 (0.918–1.420) | 0.234 | 1.106 (0.889–1.377) | 0.366 | 1.062 (0.853–1.321) | 0.591 |
| 2016 ASE/EACVI guidelines |  |  |  |  |  |  |  |  |
| Indeterminate diastolic function | 1.549 (1.146–2.094) | 0.004 | 1.340 (0.989–1.815) | 0.059 |  |  |  |  |
| DD with normal LAP | 2.212 (1.868–2.620) | <0.001 | 1.394 (1.165–1.669) | <0.001 |  |  |  |  |
| DD with indeterminate LAP | 3.905 (3.235–4.714) | <0.001 | 1.987 (1.609–2.455) | <0.001 |  |  |  |  |
| DD with elevated LAP | 5.624 (4.558–6.938) | <0.001 | 2.712 (2.164–3.399) | <0.001 |  |  |  |  |
| 2025 ASE guidelines |  |  |  |  |  |  |  |  |
| DD with normal LAP | 1.593 (1.361–1.865) | <0.001 |  |  | 1.243 (1.059–1.459) | 0.008 |  |  |
| DD with indeterminate LAP | 5.067 (3.847–6.674) | <0.001 |  |  | 1.680 (1.207–2.338) | 0.002 |  |  |
| DD with elevated LAP | 4.141 (3.423–5.010) | <0.001 |  |  | 2.202 (1.801–2.693) | <0.001 |  |  |
| DL-predicted probability of DD | 4.632 (4.000–5.364) | <0.001 |  |  |  |  | 2.673 (2.293–3.117) | <0.001 |

When we divided participants into two groups (i.e., low-risk and high-risk groups) using the DL-predicted probability value of 0.5 as the cutoff and plotted their event-free survival, we observed that a higher proportion of high-risk compared to low-risk patients reached the composite endpoint during follow-up (Figure 4). We also observed a significant separation between the survival curves of patients categorized based on the 2016 and 2025 guidelines (Figure 4, Supplemental Tables 9 and 10).

**Figure 4.**
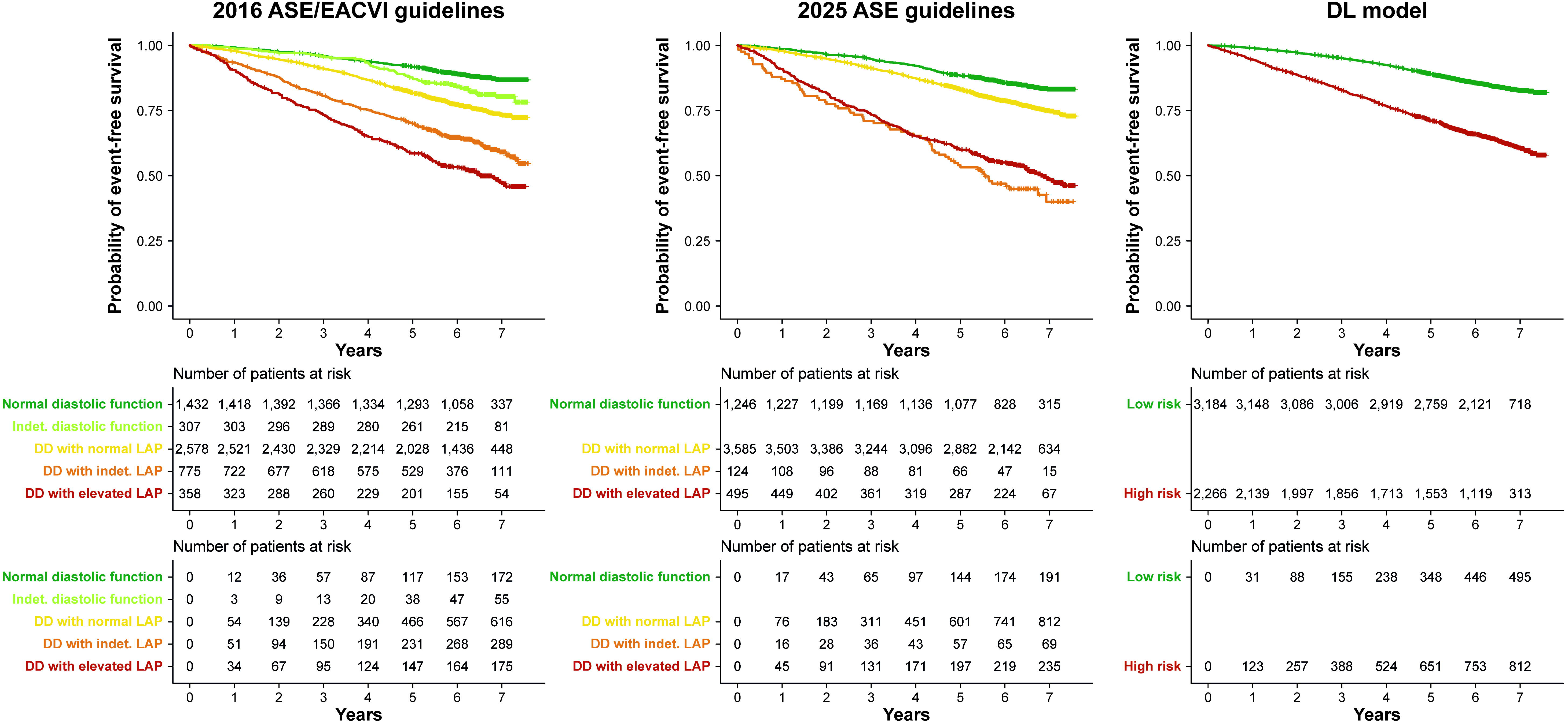
Kaplan-Meier curves showing the event-free survival of patient subgroups in the ARIC cohort The composite of HF hospitalization or all-cause death was used as the endpoint in this analysis. Abbreviation as in Figures 1 and 2.

The DL-predicted probabilities, as well as the guideline-based classifications, were also associated with HF hospitalization and all-cause death as separate endpoints in univariable and multivariable analyses (Supplemental Tables 11 and 12). Moreover, the DL model outperformed the two guidelines in predicting both of these endpoints (Supplemental Figure 3).

### Prognostic value in ARIC study participants with preserved LVEF

Of participants with available LVEF measurement at visit 5 and preserved LVEF (n=5,135 [94%], 75 [71–79] years, 42% male), 1,170 (23%) reached the composite endpoint of HF hospitalization or all-cause death over 6.5 (6.1–7.0) years. In this subgroup of ARIC participants, the DL-predicted probability of DD demonstrated the highest prognostic discrimination (C-index: 0.660 [95% CI: 0.643–0.677]), significantly outperforming the 2016 (C-index: 0.628 [95% CI: 0.613–0.643], p<0.001) and 2025 guidelines (C-index: 0.590 [95% CI: 0.576–0.604], p<0.001) and the H_2_FPEF score (C-index: 0.607 [95% CI: 0.591–0.623], p<0.001; Table 3). The 2016 guidelines also showed an improvement over the H_2_FPEF score (p=0.014; Table 3). In contrast, the 2025 ASE guidelines showed performance similar to that of the H_2_FPEF score (p=0.070; Table 3).

**Table 3.** Prognostic value of the guideline-based classifications and the DL model compared with the H_2_FPEF score in ARIC study participants with preserved LVEF.

|  | C-index | ΔC-index | P-value |
| --- | --- | --- | --- |
| **H<sub>2</sub>FPEF score** | 0.607 (0.591–0.623) |  |  |
| **2016 ASE/EACVI guidelines** | 0.628 (0.613–0.643) | +0.021 | 0.014 |
| **2025 ASE guidelines** | 0.590 (0.576–0.604) | -0.017 | 0.070 |
| **DL-predicted probability of DD** | 0.660 (0.643–0.677) | +0.053 | <0.001 |

Each row represents a univariable Cox regression model predicting the composite endpoint of HF hospitalization or all-cause death. The third column shows the difference in the C-index of the given model vs. that of the model containing the H<sub>2</sub>FPEF score only, whereas the P-value in the fourth column refers to the significance of this difference.
Abbreviations as in Table 1.

### Performance in detecting elevated LVFP

Of the 83 patients of the first hemodynamic validation cohort (61 [54–66] years, 67% male), 25 (30%) had elevated invasively measured LVFP. Invasive hemodynamic assessment was performed with LHC in 59 (71%) and RHC in 24 (29%) patients. Sixty-three (76%) patients underwent echocardiography and invasive hemodynamic assessment on the same day, and the remaining 20 (24%) underwent both tests within 72 hours (Table 1 and Supplemental Table 13). There were apparent reclassifications between the three methods, as shown by the Sankey diagram (Figure 2). Significantly fewer patients had DD with indeterminate LAP according to the 2025 guidelines than the 2016 guidelines (2 [2%] vs. 16 [19%], p<0.001). In identifying patients with elevated LVFP, the DL model exhibited an AUC of 0.879 (95% CI: 0.790–0.967), which was higher than the AUC of the 2025 guidelines (0.822 [95% CI: 0.722–0.922], p=0.041) and numerically higher, although not statistically significant, than that of the 2016 guidelines (0.812 [95% CI: 0.714–0.910], p=0.138) (Figure 5A). There was no difference between the AUCs of the 2016 and 2025 guidelines (p=0.815). The DL model outperformed both guidelines even after excluding patients with indeterminate diastolic function or DD with indeterminate LAP (Figure 5B, Table 4), patients with atrial fibrillation (Supplemental Figure 4), and those with both LARS and isovolumic relaxation time (IVRT) missing (Supplemental Figure 5).

**Figure 5.**
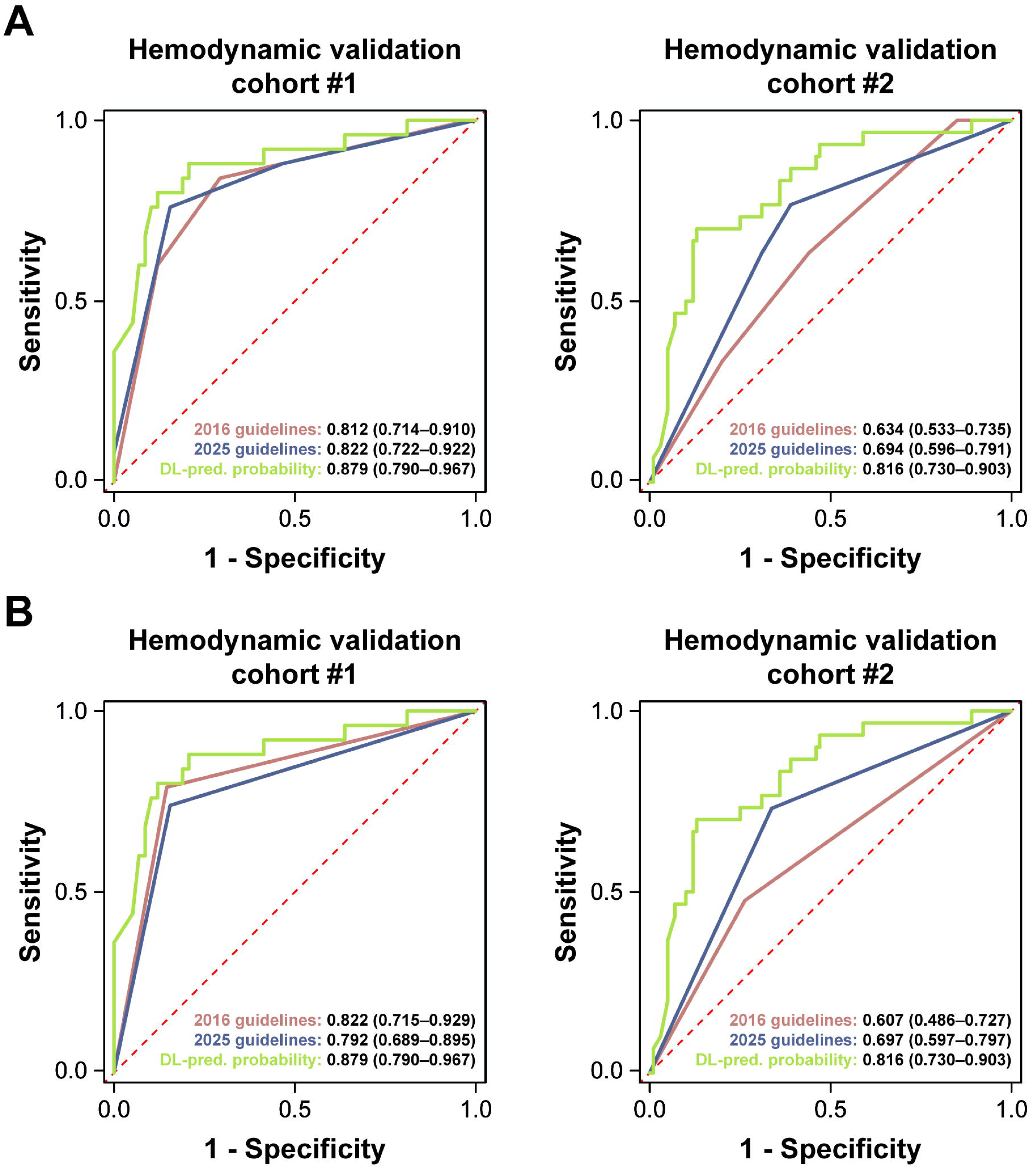
ROC curves showing the performance of the guidelines and the DL model in detecting elevated LVFP ROC analysis was performed first for all patients in the given cohort (A). Then, indeterminate groups were excluded, and ROC analyses were performed again in the remaining patients (B). Because the 2016 ASE/EACVI and 2025 ASE guidelines classified different sets of patients as indeterminate, their performance was assessed in overlapping but not identical patient subsets after excluding indeterminate cases: the 2016 guidelines were evaluated in 67 and 91 patients in the first and second hemodynamic validation cohorts, whereas the 2025 guidelines were evaluated in 79 and 118 patients, respectively. To enable the straightforward calculation of diagnostic performance metrics, both guideline-based classifications were dichotomized by merging the “Normal diastolic function” and the “DD with normal LAP” groups (as both are supposed to have normal LAP). AUCs are reported with 95% CIs. AUC – area under the receiver operating characteristic curve, LV – left ventricular, ROC – receiver operating characteristic; other abbreviations as in Figure 2.

**Table 4.** Performance of the guidelines and the DL model in detecting elevated LVFP in the first hemodynamic validation cohort (indeterminate groups excluded)

|  | 2016 ASE/EACVI guidelines | 2025 ASE guidelines | DL model |
| --- | --- | --- | --- |
| **DD with indeterminate LAP, n (%)** | 16 (19) | 2 (2) | - |
| **AUC** | 0.822 (0.715–0.929) | 0.792 (0.689–0.895) | 0.879 (0.790–0.967) |
| **Accuracy, %** | 84 | 81 | 86 |
| **Sensitivity, %** | 79 | 74 | 80 |
| **Specificity, %** | 85 | 84 | 88 |
| **PPV, %** | 68 | 65 | 74 |
| **NPV, %** | 91 | 89 | 91 |

The two guidelines classified different patients as indeterminate; therefore, their performance was evaluated in overlapping but non-identical subsets after excluding indeterminate cases: the 2016 ASE/EACVI guidelines were evaluated in 67 patients, whereas the 2025 ASE guidelines were evaluated in 79 patients. The DL model was evaluated in all patients. To enable the straightforward calculation of diagnostic performance metrics (i.e., accuracy, sensitivity, specificity, positive predictive value, and negative predictive value), both guideline-based classifications were dichotomized by merging the “Normal diastolic function” and the “DD with normal LAP” groups (as both are supposed to have normal LAP). The DL-predicted probabilities were dichotomized using a cutoff of 0.99957, optimized in this cohort by maximizing Youden’s J statistic. AUCs are reported with 95% CIs.

Of the 130 patients of the second validation cohort (66 [51–75] years, 54% male), 30 (23%) had elevated LVFP based on invasive hemodynamic assessment. Fifty (38%) patients underwent echocardiography and RHC on the same day, whereas the other 80 (62%) underwent both tests within 24 hours. The clinical and echocardiographic characteristics of this cohort are reported in Table 1 and Supplemental Table 14. The Sankey diagram showed notable reclassification in this cohort as well (Figure 2). The 2025 guidelines classified significantly fewer patients as having DD with indeterminate LAP than the 2016 guidelines (12 [9%] vs. 39 [30%], p<0.001). The DL-predicted probability detected elevated LVFP with an AUC of 0.816 (95% CI: 0.730–0.903), which was significantly higher than the AUCs of the 2016 (0.634 [95% CI: 0.533–0.735], p<0.001) and the 2025 guidelines (0.694 [95% CI: 0.596–0.791], p=0.004) (Figure 5A). The discriminatory power of the 2016 and 2025 guidelines was similar (p=0.209). The DL model also demonstrated superior performance compared with both guidelines when the indeterminate groups (Figure 5B, Table 5), patients with atrial fibrillation (Supplemental Figure 4), or those with both LARS and IVRT missing (Supplemental Figure 5) were excluded from the analysis.

**Table 5.** Performance of the guidelines and the DL model in detecting elevated LVFP in the second hemodynamic validation cohort (indeterminate groups excluded)

|  | 2016 ASE/EACVI guidelines | 2025 ASE guidelines | DL model |
| --- | --- | --- | --- |
| **Indeterminate diastolic function or DD with indeterminate LAP, n (%)** | 39 (30) | 12 (9) | - |
| **AUC** | 0.607 (0.486–0.727) | 0.697 (0.597–0.797) | 0.816 (0.730–0.903) |
| **Accuracy, %** | 68 | 68 | 77 |
| **Sensitivity, %** | 48 | 73 | 70 |
| **Specificity, %** | 74 | 66 | 79 |
| **PPV, %** | 33 | 38 | 50 |
| **NPV, %** | 84 | 90 | 89 |

The two guidelines classified different patients as indeterminate; therefore, their performance was evaluated in overlapping but non-identical subsets after excluding indeterminate cases: the 2016 ASE/EACVI guidelines were evaluated in 91 patients, whereas the 2025 ASE guidelines were evaluated in 118 patients. The DL model was evaluated in all patients. To enable the straightforward calculation of diagnostic performance metrics (i.e., accuracy, sensitivity, specificity, positive predictive value, and negative predictive value), both guideline-based classifications were dichotomized by merging the “Normal diastolic function” and the “DD with normal LAP” groups (as both are supposed to have normal LAP). To preserve the operating point derived in the first hemodynamic validation cohort while avoiding cohort-specific optimization, percentile-based operating-point transport was applied to this cohort. Accordingly, the top 32.5% of DL-predicted probabilities were classified as positive, corresponding to a cohort-specific numeric cutoff of 0.98612. AUCs are reported with 95% CIs. Abbreviations as in Tables 1 and 4.

## DISCUSSION

In this comprehensive comparative study, we evaluated the performance of the 2025 ASE guidelines, the 2016 ASE/EACVI guidelines, and a validated DL model for assessing diastolic function (Central Illustration). We leveraged the racially and demographically diverse ARIC cohort from multiple US communities and two geographically distinct hemodynamic validation cohorts with invasively measured LVFP to evaluate the generalizability and robustness of these three approaches across various populations and settings. Importantly, these cohorts reflect real-world clinical settings in which complete acquisition of Doppler and strain parameters is not uniformly achievable.

Although the 2025 guidelines markedly reduced indeterminate classifications compared with the 2016 guidelines, the DL model yielded better prognostic discrimination than both guidelines in the ARIC cohort and also outperformed the H_2_FPEF score among patients with preserved LVEF. Furthermore, the DL model demonstrated superior or similar performance in detecting elevated LVFP in the two hemodynamic validation cohorts. Notably, this performance was achieved using routinely acquired echocardiographic variables, without reliance on the full complement of advanced parameters.

### Comparison of the 2025 and 2016 guidelines

The 2016 ASE/EACVI guidelines were designed for broad clinical usability but were limited by high rates of indeterminate classifications and their moderate correlation with invasive measures of LVFP (7–11). The 2025 ASE guidelines introduced several notable improvements, including the integration of additional echocardiographic features (e.g., LARS) and the use of age-specific cutoff values (5). They also substantially reduce the proportion of indeterminate cases, as recently demonstrated by Lababidi et al. (12). Consistent with their findings, we observed a marked reduction in indeterminate classifications, from 20% to 2% in the ARIC cohort, from 19% to 2% in the first hemodynamic validation cohort, and from 30% to 9% in the second hemodynamic validation cohort, despite missing pulmonary vein systolic and diastolic velocities (S/D) and limited availability of LARS, reflecting real-world variability in data acquisition.

Nevertheless, despite the advantages noted above, the 2025 guidelines did not surpass the 2016 guidelines in prognostic or diagnostic performance in our analysis. These findings contrast with those of Lababidi et al. (12), likely reflecting differences in patient selection (e.g., exclusion of patients with persistent or permanent atrial fibrillation) and analytic approach. Specifically, they used the McNemar test to compare paired classification accuracies between the guidelines, which required excluding all cases classified as indeterminate by either guideline version (12). While this approach emphasizes performance in definitively classifiable patients, it may overlook the real-world clinical burden of indeterminate classifications. Therefore, we reported two sets of performance metrics: one that included all patients, with indeterminate results treated as non-conclusive, and another that excluded indeterminate cases separately for each guideline version. Importantly, our findings align with those of Harada et al. (13), who reported limited discrimination of the 2025 guidelines despite fewer indeterminate cases. Taken together, these results suggest that improvements in classification do not necessarily translate into higher accuracy, highlighting limitations of rule-based approaches.

### Comparison of the DL model and expert guidelines

In contrast to rule-based algorithms, the DL model demonstrated consistently strong performance across cohorts. The DL-predicted probability of DD provided additive value over NT-proBNP, confirming its capability to capture risk and disease severity beyond conventional biomarkers. The DL model also outperformed the H_2_FPEF score in patients with preserved LVEF, supporting its role as a complementary tool for risk stratification.

A salient feature of the DL model meriting recapitulation is that it was trained to capture complex, nonlinear relationships among multiple echocardiographic parameters using a hybrid approach combining unsupervised learning with supervised classification (15). This approach enabled the model to integrate echocardiographic parameters within a multivariable probabilistic framework and to successfully overcome key limitations of traditional threshold-based algorithms, including age-dependent shifts in reference ranges and challenges caused by missing data. Prior work demonstrated that the model encodes physiologically meaningful patterns (29), supporting its robustness and potential clinical applicability (15).

### Clinical implications and future directions

The findings have several clinical implications. First, HFpEF diagnosis remains challenging and often requires invasive hemodynamic examinations, leading to delays and missed opportunities. A probabilistic, data-driven model may provide a more individualized assessment than rule-based algorithms and enable earlier identification of high-risk patients.

Second, the emergence of several new HFpEF therapies, including sodium-glucose cotransporter-2 inhibitors (30), glucagon-like peptide-1 receptor agonists (31), and finerenone (32), underscores the importance of early diagnosis, as improved risk stratification may facilitate timely initiation of these disease-modifying therapies and improve outcomes.

Third, the DL model may support clinical trials and longitudinal research by enabling standardized phenotyping. Fourth, its compatibility with routinely acquired echocardiographic parameters makes the model well-suited for real-world implementation through integration into existing reporting software solutions or electronic health record systems to assist clinical decision-making.

### Limitations

Despite its strengths, the present study has several limitations. First, some parameters recommended in the 2025 guidelines (e.g., LARS, IVRT, pulmonary vein S/D) were not consistently available. Notably, other recent studies evaluating the 2025 guidelines, including the one by Harada et al., similarly lacked pulmonary vein Doppler signals due to feasibility constraints (13). Furthermore, even in the initial validation study by Lababidi et al., pulmonary vein S/D and atrial velocity duration were feasible in only 48% and 38% cases, respectively (12). Therefore, missing data reflect real-world practice rather than a study-specific limitation. Importantly, despite the incomplete availability of these parameters, the DL model exhibited robust diagnostic and prognostic performance using only routinely acquired variables.

Second, the DL model’s performance may partly reflect its continuous output compared with categorical classifications. However, superiority persisted even after dichotomizing its predictions (Supplemental Results).

Third, prospective implementation of the DL model remain to be established in terms of patient outcomes, workflow efficiency, or treatment decisions. Integration into mobile and cloud-based platforms could help bridge this gap by facilitating prospective implementation and enabling remote HF screening and triage, particularly in resource-limited settings.

Fourth, despite previous foundational work showing that the DL model can identify transparent phenogroups (15), clinicians may still perceive it as a “black box”.

Lastly, the sample size of the hemodynamic validation cohorts was modest, and only resting hemodynamics were available. Further validation in larger and more diverse cohorts is needed.

## CONCLUSIONS

The DL model for assessing LV diastolic dysfunction demonstrated the potential for improved diagnostic accuracy and prognostic discrimination compared to both the 2025 ASE and 2016 ASE/EACVI guidelines. Importantly, the DL model achieved this performance using only routinely available echocardiographic variables, suggesting that data-driven approaches capable of extracting latent physiologic information may provide a scalable framework for diastolic function assessment, particularly in settings where advanced parameters are not consistently obtainable. Future prospective studies are needed to evaluate these methods in head-to-head comparisons, particularly in real-world settings, to refine risk stratification and support more personalized patient selection for emerging targeted therapies as cardiovascular care advances toward precision medicine.

## Data Availability

The data used in this study are not publicly available because of Institutional Review Board (IRB) restrictions and participant privacy considerations. De-identified analytical results and supporting materials can be made available from the corresponding author upon reasonable request, subject to applicable ethical and institutional approvals.

---

**Central Illustration.**
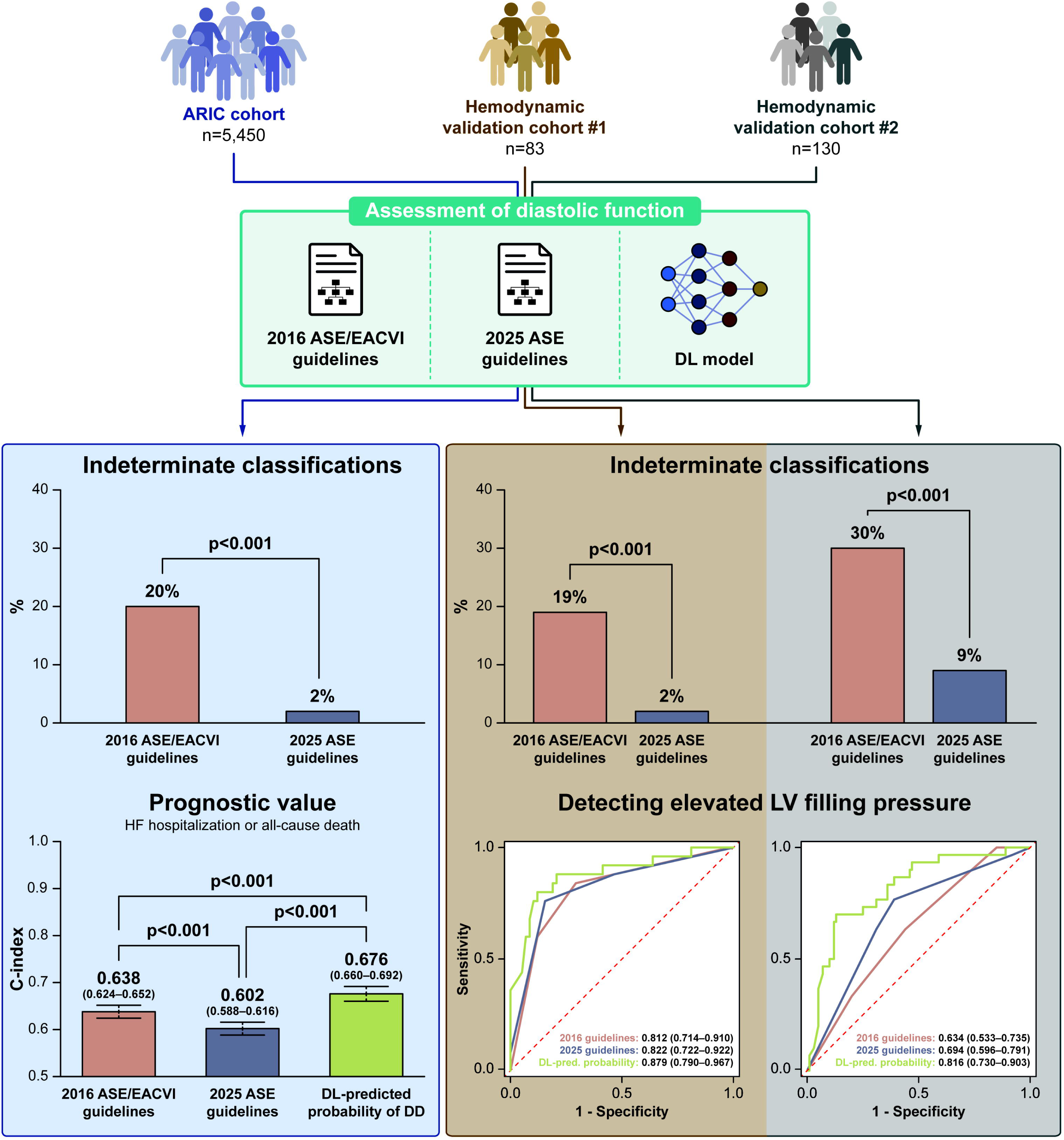
Comparison of the expert guidelines with AI-driven echocardiographic assessment of diastolic function In this study, we compared the diagnostic and prognostic performance of the newly released 2025 ASE guidelines, the earlier 2016 ASE/EACVI guidelines, and a rigorously validated DL model for assessing LV diastolic function. Although the 2025 guidelines markedly reduced indeterminate classifications in all three cohorts compared with the 2016 guidelines, they demonstrated lower prognostic value and similar diagnostic performance for detecting elevated LVFP in the hemodynamic validation cohorts. The DL model yielded better prognostic discrimination than both guidelines in the ARIC cohort and showed superior performance in detecting elevated LVFP. AI – artificial intelligence, ARIC – Atherosclerosis Risk In Communities, ASE – American Society of Echocardiography, DD – diastolic dysfunction, DL – deep learning, EACVI – European Association of Cardiovascular Imaging, LV – left ventricular, LVFP – left ventricular filling pressure

