## Supplemental Material for "Comparison of the Expert Guidelines With Artificial Intelligence-Driven Echocardiographic Assessment of Diastolic Function"

#### SUPPLEMENTAL METHODS

##### Echocardiographic protocol in the ARIC cohort study

Echocardiographic examinations were performed by experienced echocardiographers at each field center using commercially available ultrasound systems (iE33, Philips Medical Systems with Vision 2011). The protocol included 2D, color Doppler, spectral Doppler, and tissue Doppler imaging. All measurements and evaluations were conducted at the Atherosclerosis Risk in Communities (ARIC) Echocardiography Reading Center (Brigham and Women's Cardiac Imaging Core Laboratory, Boston, MA). All analyses were over-read (blinded to study participants' clinical information) and were approved by board-certified echocardiographers.

##### Calculation of the $H_2FPEF$ score

$H_2FPEF$ score was calculated according to the original published algorithm (1), which incorporates the following six clinical and echocardiographic variables: body mass index >30 kg/m² (2 points); use of ≥2 antihypertensive medications (1 point); paroxysmal or persistent atrial fibrillation (3 points); Doppler echocardiography-estimated pulmonary artery systolic pressure >35 mmHg (1 point); age >60 years (1 point); and ratio of early mitral inflow velocity and early diastolic mitral annular velocity at septal position (E/e') >9 (1 point). Each component was assigned a point value based on the original weighting scheme, resulting in total scores ranging from 0 to 9, with higher scores indicating a greater likelihood of heart failure with preserved ejection fraction (EF).

**Development of the DL model for assessing diastolic function**

To develop a deep learning (DL) model for assessing diastolic function, topological data analysis (TDA) was first performed using the Ayasdi Workbench (version 7.9, Ayasdi Inc., Palo Alto, California) to simultaneously evaluate nine echocardiographic features (i.e., left ventricular mass index [LVMi], left ventricular EF [LVEF], early mitral inflow velocity [E], late mitral inflow velocity [A], E/A, septal e', septal E/e', left atrial volume index [LAVi], and tricuspid regurgitation peak velocity [TRV]) of 1,242 patients with varying degrees of systolic and diastolic dysfunction (2,3). TDA applies the tools of shape analysis to identify and connect highly similar data points (e.g., patients) in a multi-dimensional space and then plots the identified connections as a two-dimensional similarity network (4). The generated network consists of nodes (representing collections of similar data points) connected by edges (i.e., lines between two nodes) if they have at least one data point in common. In the analyzed dataset, TDA yielded a circular similarity network that was divided into two parts based on the differences in clinical characteristics and outcomes of patients located in its different regions: one comprising patients with low risk and the other encompassing patients with a high risk of diastolic dysfunction (DD). These labels were assigned to each of the 1,242 patients in the similarity network and were used to train classifiers in a subsequent supervised learning phase, enabling the assessment of diastolic function in new, previously unseen patients.

For training and evaluation of the classifiers, BigML (BigML Inc., Corvallis, OR, http://bigml.com), a cloud-based machine learning platform, was utilized. After splitting the cohort into training and test sets in an 80:20 ratio, various classifiers (i.e., decision trees, ensembles of decision trees, logistic regression models, and deep neural networks) were trained on the training set (with cross-validation) and subsequently evaluated on the test set. Missing values were handled natively by the learning algorithm: missing values in categorical features were treated as a distinct input category, while missing numeric values were explicitly modeled rather than imputed or excluded, both during training and inference, using BigML's default configuration. Model selection and hyperparameter tuning were performed using the OptiML optimization process, which leverages Bayesian optimization to evaluate hundreds of supervised models and identify the best-performing ones (5). These models can then be used individually or combined into a so-called *fusion model*.

The final DL model is a *fusion model* that combines the seven best-performing deep neural networks and averages their predictions (3). In this model, the most important feature for predicting DD is e', followed by E/e', LVMi, LVEF, LAVi, TRV, E, A, and E/A in decreasing order of importance. The model has been externally validated in multiple cohorts, showing that higher DL-predicted DD probability values were associated with elevated LV filling pressures (LVFP), increased neurohormonal activation, lower exercise capacity, and a higher incidence of adverse cardiac events during follow-up (3). The DL model is publicly available at https://wvu-model.herokuapp.com. A more detailed description of model development and evaluation has been published previously (2,3,6).

**Dichotomizing the output of the DL model**

To verify that the DL model's superior performance over the guidelines was not solely attributable to its ability to generate continuous risk estimates, in contrast to the categorical nature of the guideline-based classifications, C-indices and areas under the receiver operating characteristic curve (AUCs) were also calculated and compared after dichotomizing the DL model's output. In the ARIC cohort, the default 0.5 threshold was used to dichotomize the DL-predicted probabilities. Given that the DL model was primarily trained for prognostication, a cutoff optimized for identifying elevated LVFP was applied instead of the default 0.5 threshold to dichotomize the predicted probabilities in the hemodynamic validation cohorts. In the first hemodynamic validation cohort, the classification threshold was selected by maximizing

Youden’s J statistic, yielding an optimal cutoff of 0.99957 and a predicted-positive rate of 32.5%. To preserve the operating point derived in the first cohort while avoiding cohort-specific optimization, percentile-based operating-point transport was applied to the second hemodynamic validation cohort (7). Accordingly, the top 32.5% of DL-predicted probabilities were classified as positive in that cohort, corresponding to a cohort-specific numeric cutoff of 0.98612. To ensure a fair comparison, the outputs of both guideline-based classifications were also dichotomized by combining all categories except for “DD with elevated LAP”.

### SUPPLEMENTAL RESULTS

**Results using the dichotomized output of the DL model**

The DL model demonstrated superior prognostic power over both guidelines in univariable Cox regression even when its predicted probabilities were dichtomized, with a C-index of 0.627 (95% CI: 0.614–0.640) vs. 0.543 (95% CI: 0.534–0.552) for the 2016 guidelines and 0.556 (95% CI: 0.546–0.566) for the 2025 guidelines ($p<0.001$ for both comparisons).

In the first hemodynamic validation cohort, the dichotomized output of the DL model exhibited an AUC of 0.840 (95% CI: 0.749–0.930) in identifying patients with elevated LVFP, which was higher than the AUC of the 2025 guidelines (0.728 [95% CI: 0.621–0.835], $p=0.041$) and was numerically higher than that of the 2016 guidelines, although the latter difference did not reach statistical significance (0.740 [95% CI: 0.633–0.846], $p=0.066$).

In the second hemodynamic validation cohort, the dichotomized output of the DL model detected elevated LVFP with an AUC of 0.785 (95% CI: 0.695–0.875), which was significantly higher than the AUCs of the 2016 (0.567 [95% CI: 0.472–0.661], $p<0.001$) and the 2025 guidelines (0.662 [95% CI: 0.563–0.761], $p=0.012$).

**Supplemental Table 1.** Availability of the echocardiographic parameters used for LV diastolic function assessment in the three cohorts

| | ARIC cohort | Hemodynamic validation cohort #1 | Hemodynamic validation cohort #2 |
|---|---|---|---|
| **LVEF** | 97 | 100 | 100 |
| **LVMi** | 99 | 94 | 98 |
| **E** | 100 | 100 | 100 |
| **E-DT** | 100 | 99 | 99 |
| **A** | 96 | 94 | 83 |
| **E/A** | 96 | 94 | 83 |
| **IVRT** | 0 | 75 | 42 |
| **e’ (septal)** | 100 | 100 | 100 |
| **E/e’ (septal)** | 100 | 100 | 100 |
| **e’ (lateral)** | 100 | 98 | 60 |
| **E/e’ (lateral)** | 100 | 98 | 60 |
| **e' (average)** | 100 | 98 | 60 |
| **E/e’ (average)** | 100 | 98 | 60 |
| **LAVi** | 99 | 96 | 83 |
| **LARS** | 0 | 78 | 95 |
| **TRV** | 58 | 53 | 96 |
| **PASP** | 0 | 37 | 52 |
| **Pulmonary vein S/D** | 97 | 100 | 100 |

Cell values are the percentages denoting the availability of each echocardiographic parameter. Cells are color-coded based on the percentage value, with green indicating high availability and red indicating low availability.

A – late mitral inflow velocity, ARIC – Atherosclerosis Risk in Communities study, D – pulmonary vein diastolic velocity, E – early mitral inflow velocity, E-DT – E-wave deceleration time, e’ – early diastolic mitral annular velocity, IVRT – isovolumic relaxation time, LARS – left atrial reservoir strain, LAVi – left atrial volume index, LV – left ventricular, LVEF – left ventricular ejection fraction, LVMi – left ventricular mass index, PASP – pulmonary artery systolic pressure estimated by echocardiography, S – pulmonary vein systolic velocity, TRV – tricuspid regurgitation peak velocity

**Supplemental Table 2.** Proposed Requirements for Cardiovascular Imaging-Related Multimodal-AI Evaluation (PRIME 2.0) checklist

| Checklist item | Manuscript section/figure/table |
|---|---|
| **1. Designing an AI study in cardiovascular imaging** | |
| 1.1 Appropriateness of applying AI | |
| Describe the need for applying AI | Introduction |
| Determine the appropriateness of applying AI | Introduction |
| 1.2 Study objectives, input data type, and prediction target | |
| Explain the AI task and the likely deployment context | Introduction<br>Discussion<br>Also explained previously in the paper reporting the development and initial validation of the DL model: https://doi.org/10.1016/j.jcmg.2021.04.010 |
| Describe the input data, number of training/test examples | Methods – Validation cohort for predicting clinical outcomes<br>Methods – Validation cohorts for detecting elevated LVFP<br>Figure 1, Table 1<br>Supplemental Methods – Development of the DL model for assessing diastolic function<br>Also described previously in the paper reporting the development and initial validation of the DL model: https://doi.org/10.1016/j.jcmg.2021.04.010 |
| Specify model supervision type | Methods – DL-based assessment of diastolic function<br>Supplemental Methods – Development of the DL model for assessing diastolic function<br>Discussion – Comparison of the DL model and expert guidelines<br>Also described previously in the paper reporting the development and initial validation of the DL model: https://doi.org/10.1016/j.jcmg.2021.04.010 |
| Describe the nature of the model's output and what it represents | Methods – DL-based assessment of diastolic function<br>Supplemental Methods – Development of the DL model for assessing diastolic function<br>Also described previously in the paper reporting the development and initial validation of the DL model: https://doi.org/10.1016/j.jcmg.2021.04.010 |
| 1.3 Design of the AI study | |
| Describe the study design | External validation of a pre-existing DL model using data previously collected in prospective studies and registries.<br>Central Illustration |

| Checklist item | Manuscript section/figure/table |
| --- | --- |
| Describe data origin | Methods – Validation cohort for predicting clinical outcomes<br>Methods – Validation cohorts for detecting elevated LVFP |
| Describe whether impact analysis was included | Impact analysis was not performed. |
| **2. Data format and preprocessing** | |
| 2.1 Data format | |
| Describe the technical details of the data acquisition | Methods – Validation cohort for predicting clinical outcomes<br>Methods – Validation cohorts for detecting elevated LVFP<br>Supplemental Methods – Echocardiographic protocol in the ARIC cohort study<br>Also described previously in the paper reporting the development and initial validation of the DL model: https://doi.org/10.1016/j.jcmg.2021.04.010 |
| Describe the technical details of the data format | Only tabular data was used in this study. |
| 2.2 Clinical characteristics of the study cohort | |
| Present the age, sex, and race distributions of the cohorts | Table 1<br>Supplemental Tables 3, 13, and 14 |
| Summarize key clinical and imaging characteristics of the cohorts | Table 1<br>Supplemental Tables 3, 13, and 14 |
| Compare summary statistics of cases and controls | Supplemental Tables 3, 13, and 14 |
| 2.3 Steps of data preprocessing | |
| Describe how data were cleaned, made uniform, and consistent | Methods – Validation cohort for predicting clinical outcomes<br>Methods – Validation cohorts for detecting elevated LVFP<br>Supplemental Methods – Echocardiographic protocol in the ARIC cohort study<br>Also described previously in the paper reporting the development and initial validation of the DL model: https://doi.org/10.1016/j.jcmg.2021.04.010 |
| Describe data harmonization techniques (if applicable) | N/A |
| Provide details on missing values and imputation methods | Supplemental Tables 1, 3, 4, 13, and 14 |
| Describe processes for handling outliers | Supplemental Methods – Development of the DL model for assessing diastolic function<br>Also described previously in the paper reporting the development and initial validation of the DL model: https://doi.org/10.1016/j.jcmg.2021.04.010 |

| Checklist item | Manuscript section/figure/table |
|---|---|
| Describe whether class imbalance exists | Results – Prognostic value in the entire ARIC cohort<br>Results – Prognostic value in ARIC study participants with preserved LVEF<br>Results – Performance in detecting elevated LVFP |
| 2.4 Feature engineering and feature selection | |
| Describe applied feature engineering techniques | N/A |
| Describe applied feature selection techniques | Described previously in the paper reporting the development and initial validation of the DL model: https://doi.org/10.1016/j.jcmg.2021.04.010 |
| **3. Selection of AI methods and applications** | |
| 3.1 Selecting appropriate AI methods and applications | |
| Clearly define data composition (structured/unstructured) | Supplemental Methods – Development of the DL model for assessing diastolic function<br>Also described previously in the paper reporting the development and initial validation of the DL model: https://doi.org/10.1016/j.jcmg.2021.04.010 |
| 3.2–3.4 Training strategies | |
| Describe the AI method used, with rationale for clinical task fit | Supplemental Methods – Development of the DL model for assessing diastolic function<br>Also described previously in the paper reporting the development and initial validation of the DL model: https://doi.org/10.1016/j.jcmg.2021.04.010 |
| 3.5 Solving clinical problems | Supplemental Methods – Development of the DL model for assessing diastolic function<br>Also described previously in the paper reporting the development and initial validation of the DL model: https://doi.org/10.1016/j.jcmg.2021.04.010 |
| **4. Model assessment** | |
| 4.1 Importance of model assessment | |
| Describe the evaluation approach and how it addresses the clinical question | Methods – Statistical analysis<br>Supplemental Methods – Development of the DL model for assessing diastolic function<br>Also described previously in the paper reporting the development and initial validation of the DL model: https://doi.org/10.1016/j.jcmg.2021.04.010 |

| Checklist item | Manuscript section/figure/table |
|---|---|
| 4.2 Technical performance metrics | |
| Report relevant performance metrics | Results – Prognostic value in the entire ARIC cohort<br>Results – Prognostic value in ARIC study participants with preserved LVEF<br>Results – Performance in detecting elevated LVFP<br>Tables 3, 4, and 5<br>Figures 3 and 5<br>Supplemental Tables 6, 7, and 8<br>Supplemental Figures 1, 2, 3, 4, and 5 |
| Justify metric selection based on task characteristics | Methods – Statistical analysis |
| Describe manual annotation process, reference-standard prep, and observer variability | Methods – Validation cohort for predicting clinical outcomes<br>Methods – Validation cohorts for detecting elevated LVFP<br>Supplemental Methods – Echocardiographic protocol in the ARIC cohort study |
| 4.3–4.4 Robustness, generalizability, and evaluating data quality | |
| Evaluate model robustness to external variations | Results – Prognostic value in the entire ARIC cohort<br>Results – Prognostic value in ARIC study participants with preserved LVEF<br>Results – Performance in detecting elevated LVFP<br>Tables 3, 4, and 5<br>Figures 3 and 5<br>Supplemental Tables 6, 7, and 8<br>Supplemental Figures 1, 2, 3, 4, and 5<br>Also assessed previously in these papers:<br>https://doi.org/10.1016/j.jcmg.2021.04.010<br>https://doi.org/10.1016/j.jcmg.2024.07.017<br>https://doi.org/10.1093/ehjci/jeae037 |
| Assess performance across clinical subpopulations and sociodemographic subgroups | |
| 4.5 Identifying features learned by the model | |
| Describe interpretability/explainability methods | Supplemental Methods – Development of the DL model for assessing diastolic function<br>Also described previously in the paper reporting the development and initial validation of the DL model:<br>https://doi.org/10.1016/j.jcmg.2021.04.010 |
| Quantify and discuss uncertainty | N/A |
| **5. Clinical evaluation** | |
| 5.1 Importance of clinical evaluation | |
| Describe potential clinical impact of misclassification | Misclassification could lead to delayed diagnosis, inappropriate risk stratification, and suboptimal patient management. |

| Checklist item | Manuscript section/figure/table |
|---|---|
| 5.2 Clinical utility metrics | |
| Define clinical utility of the AI system | Results – Prognostic value in the entire ARIC cohort<br>Results – Prognostic value in ARIC study participants with preserved LVEF<br>Results – Performance in detecting elevated LVFP<br>Tables 3, 4, and 5, Figures 3 and 5<br>Supplemental Tables 6, 7, and 8<br>Supplemental Figures 1, 2, 3, 4, and 5<br>Discussion – Clinical implications and future directions |
| Describe cost-effectiveness analysis | Cost-effectiveness analysis was not performed. |
| 5.3 Clinical validation | |
| Provide evidence of clinical validation | Results – Prognostic value in the entire ARIC cohort<br>Results – Prognostic value in ARIC study participants with preserved LVEF<br>Results – Performance in detecting elevated LVFP<br>Tables 3, 4, and 5, Figures 3 and 5<br>Supplemental Tables 6, 7, and 8<br>Supplemental Figures 1, 2, 3, 4, and 5 |
| 5.4 Continuous monitoring | |
| Outline plans for post-deployment monitoring | The DL model's performance will be assessed in additional validation cohorts. If performance degradation is detected, the precise cause will be identified, and the model will be refined (e.g., by updating training data [retraining], model parameter values [recalibrating], or improving the modeling structure or training method). |
| **6. Best practices for replicability** | |
| 6.1 Importance of transparency and open science principles | |
| Ensure data sharing follows FAIR (findability, accessibility, interoperability, and reusability principles | Data could not be shared due to proprietary and institutional restrictions. Access to the ARIC dataset can be requested at the following website: https://biolincc.nhlbi.nih.gov/studies/aric/ |
| Report on training data representativeness | Reported previously in the paper describing the development and initial validation of the DL model: https://doi.org/10.1016/j.jcmg.2021.04.010 |
| 6.2 Ensuring technical reproducibility | |
| Report results for the final model and training process | Supplemental Methods – Development of the DL model for assessing diastolic function<br>Also reported previously in the paper describing the development and initial validation of the DL model: https://doi.org/10.1016/j.jcmg.2021.04.010 |
| Describe sources of randomness in training | |
| Report random seeds (if applicable) | |
| Describe hardware setup | |
| Evaluate uncertainty from randomness | |

| Checklist item | Manuscript section/figure/table |
|---|---|
| Justify if the source code, model weights, or datasets are not shared | Although these are not publicly available due to proprietary and institutional restrictions, the DL model is accessible online: https://wvu-model.herokuapp.com |
| 6.3 Specific considerations for reproducing large language model and generative AI studies | N/A |
| **7. Reporting of limitations, biases, and alternatives** | |
| 7.1 Acknowledging study and model limitations | |
| Discuss key limitations | Discussion – Limitations |
| Report sensitivity analyses and model-specific issues | Results – Prognostic value in the entire ARIC cohort<br>Results – Prognostic value in ARIC study participants with preserved LVEF<br>Results – Performance in detecting elevated LVFP<br>Tables 3, 4, and 5, Figures 3 and 5<br>Supplemental Tables 6, 7, and 8<br>Supplemental Figures 1, 2, 3, 4, and 5<br>Also reported previously in the paper describing the development and initial validation of the DL model: https://doi.org/10.1016/j.jcmg.2021.04.010 |
| 7.2 Discussing study strengths | |
| Articulate strengths | Discussion |
| 7.3 Reporting on bias and fairness assessment | |
| Report stratified performance metrics for demographic subgroups | Results – Prognostic value in the entire ARIC cohort<br>Results – Prognostic value in ARIC study participants with preserved LVEF<br>Results – Performance in detecting elevated LVFP<br>Tables 3, 4, and 5, Figures 3 and 5<br>Supplemental Tables 6, 7, and 8<br>Supplemental Figures 1, 2, 3, 4, and 5 |
| Describe bias mitigation strategies and residual bias | Discussion – Limitations |
| Acknowledge fairness evaluation limitations | Discussion – Limitations |
| 7.4 Contextualizing with alternative methods | |
| Benchmark against clinical standards/traditional risk scores | The objective of this study was to compare the diagnostic and prognostic performance of the DL model with the 2016 ASE/EACVI and 2025 ASE guidelines. |
| Justify complex AI models over simpler alternatives | Justified previously in the paper reporting the development and initial validation of the DL model: https://doi.org/10.1016/j.jcmg.2021.04.010 |
| Discuss complementary methods for validation | N/A |

AI – artificial intelligence, ASE – American Society of Echocardiography, DL – deep learning, EACVI – European Association of Cardiovascular Imaging, LVFP – left ventricular filling pressure; other abbreviations as in Supplemental Table 1.

**Supplemental Table 3.** Clinical and echocardiographic characteristics of patients reaching and not reaching the composite endpoint of HF hospitalization or all-cause death in the ARIC cohort

| | **Missing** n (%) | **All participants** n=5,450 | **Participants reaching the composite endpoint** n=1,320 | **Participant not reaching the composite endpoint** n=4,130 | **P-value** |
|---|---|---|---|---|---|
| **Demographics, vitals, risk factors** | | | | | |
| Age, years | 0 (0) | 75 (71–79) | 78 (73–83) | 74 (71–78) | <0.001 |
| Male sex | 0 (0) | 2,326 (43) | 681 (52) | 1,645 (40) | <0.001 |
| Black race | 0 (0) | 1,049 (19) | 236 (18) | 813 (20) | 0.159 |
| BMI, kg/m$^2$ | 31 (1) | 27.9 (24.9–31.5) | 27.9 (24.7–31.9) | 27.9 (24.9–31.4) | 0.851 |
| BSA, m$^2$ | 7 (0) | 1.86 (1.70–2.01) | 1.88 (1.70–2.03) | 1.85 (1.70–2.01) | 0.093 |
| SBP, mmHg | 14 (0) | 129 (118–141) | 131 (119–143) | 128 (118–140) | <0.001 |
| DBP, mmHg | 14 (0) | 66 (59–74) | 64 (57–72) | 67 (60–74) | <0.001 |
| HR, 1/min | 14 (0) | 64 (57–71) | 65 (58–73) | 64 (57–71) | <0.001 |
| Hypertension | 0 (0) | 4,391 (81) | 1,165 (88) | 3,226 (78) | <0.001 |
| Diabetes | 0 (0) | 2,004 (37) | 626 (47) | 1,378 (33) | <0.001 |
| Atrial fibrillation | 0 (0) | 166 (3) | 84 (6) | 82 (2) | <0.001 |
| Chronic kidney disease | 0 (0) | 1,500 (28) | 522 (40) | 978 (24) | <0.001 |
| History of HF | 0 (0) | 908 (17) | 434 (33) | 474 (11) | <0.001 |
| History of CHD | 0 (0) | 1,048 (19) | 425 (32) | 623 (15) | <0.001 |
| History of non-cardiac PH | 0 (0) | 120 (2) | 73 (6) | 47 (1) | <0.001 |
| History of stroke | 0 (0) | 208 (4) | 91 (7) | 117 (3) | <0.001 |
| **Laboratory results** | | | | | |
| Hemoglobin, g/dL | 152 (3) | 13.3 (12.4–14.2) | 13.3 (12.4–14.2) | 13.3 (12.4–14.2) | 0.777 |
| Creatinine, mg/dL | 25 (0) | 0.92 (0.78–1.10) | 0.99 (0.84–1.24) | 0.90 (0.77–1.06) | <0.001 |
| GFR, mL/min/1.73m$^2$ | 25 (0) | 71 (59–83) | 65 (52–79) | 72 (61–83) | <0.001 |
| NT-proBNP, pg/mL | 163 (3) | 134 (69–264) | 247 (122–590) | 111 (60–210) | <0.001 |
| **Echocardiographic parameters** | | | | | |
| IVSd, mm | 36 (1) | 10 (9–11) | 11 (10–12) | 1 (9–11) | <0.001 |
| LVPWd, mm | 27 (0) | 9 (8–10) | 9 (9–11) | 9 (8–10) | <0.001 |
| LVIDd, mm | 37 (1) | 44 (41–47) | 45 (41–49) | 43 (40–47) | <0.001 |
| LVIDs, mm | 32 (1) | 26 (23–29) | 27 (24–32) | 26 (23–29) | <0.001 |
| LVMi, g/m$^2$ | 46 (1) | 76.4 (66.1–89.1) | 83.6 (71.8–102.4) | 74.3 (65.0–86.2) | <0.001 |
| LVEDVi, mL/m$^2$ | 166 (3) | 42.4 (36.4–49.7) | 44.5 (37.9–53.4) | 41.8 (36.0–48.8) | <0.001 |
| LVESVi, mL/m$^2$ | 166 (3) | 14.4 (11.6–17.9) | 15.6 (12.4–20.5) | 14.1 (11.5–17.2) | <0.001 |
| LVEF, % | 160 (3) | 65.8 (61.9–69.2) | 64.3 (59.4–68.2) | 66 (62.5–69.5) | <0.001 |
| LAVi, mL/m$^2$ | 54 (1) | 24.7 (20.3–30.2) | 27.8 (22.4–34.6) | 24.0 (19.7–28.9) | <0.001 |
| Moderate or greater AS | 0 (0) | 40 (1) | 24 (2) | 16 (0) | <0.001 |
| Moderate or greater AR | 0 (0) | 22 (0) | 9 (1) | 13 (0) | 0.114 |
| Moderate MR | 0 (0) | 98 (2) | 43 (3) | 55 (1) | <0.001 |
| E, cm/s | 0 (0) | 65 (54–78) | 68 (55–84) | 64 (54–76) | <0.001 |
| E-DT, ms | 19 (0) | 200 (173–230) | 197 (170–229) | 203 (177–230) | <0.001 |
| A, cm/s | 229 (4) | 79 (67–91) | 82 (69–96) | 78 (67–90) | <0.001 |
| E/A | 229 (4) | 0.8 (0.7–1.0) | 0.8 (0.6–1.0) | 0.8 (0.7–1.0) | 0.050 |

**Supplemental Table 3.** Continued

| | **Missing** n (%) | **All participants** n=5,450 | **Participants reaching the composite endpoint** n=1,320 | **Participant not reaching the composite endpoint** n=4,130 | **P-value** |
|---|---|---|---|---|---|
| e' (septal), cm/s | 0 (0) | 5.5 (4.7–6.5) | 5.1 (4.4–6.1) | 5.6 (4.8–6.6) | <0.001 |
| E/e' (septal) | 0 (0) | 11.6 (9.5–14.4) | 12.9 (10.2–16.5) | 11.3 (9.3–13.8) | <0.001 |
| e' (lateral), cm/s | 15 (0) | 6.8 (5.6–8.2) | 6.5 (5.2–8.0) | 6.9 (5.7–8.3) | <0.001 |
| E/e' (lateral) | 15 (0) | 9.5 (7.6–12.0) | 10.3 (8.0–13.3) | 9.3 (7.5–11.6) | <0.001 |
| e' (average), cm/s | 15 (0) | 6.2 (5.3–7.3) | 5.9 (4.9–7.0) | 6.3 (5.4–7.4) | <0.001 |
| E/e' (average) | 15 (0) | 10.6 (8.7–13.1) | 11.8 (9.4–14.8) | 10.3 (8.5–12.6) | <0.001 |
| TRV, m/s | 2,273 (42) | 2.36 (2.18–2.55) | 2.45 (2.24–2.68) | 2.34 (2.17–2.52) | <0.001 |
| 2016 ASE/EACVI guidelines | 0 (0) | | | | |
| Normal diastolic function | | 1,432 (26) | 172 (12) | 1,260 (31) | |
| Indet. diastolic function | | 307 (6) | 56 (4) | 251 (6) | |
| DD with normal LAP | | 2,578 (47) | 621 (47) | 1,957 (47) | |
| DD with indeterminate LAP | | 775 (14) | 294 (22) | 481 (12) | |
| DD with elevated LAP | | 358 (7) | 177 (13) | 181 (4) | <0.001 |
| 2025 ASE guidelines | 0 (0) | | | | |
| Normal diastolic function | | 1,246 (23) | 191 (14) | 1,055 (26) | |
| DD with normal LAP | | 3,585 (66) | 822 (62) | 2,763 (67) | |
| DD with indeterminate LAP | | 124 (2) | 69 (5) | 55 (1) | |
| DD with elevated LAP | | 495 (9) | 238 (18) | 257 (6) | <0.001 |
| DL-predicted probability of DD | 0 (0) | 0.30 (0.04–0.86) | 0.80 (0.14–0.97) | 0.18 (0.04–0.74) | <0.001 |

Continuous variables are presented as median (interquartile range) and categorical variables as n (%). Comparisons between patients reaching and not reaching the composite endpoint were performed using unpaired Student's t-test or Mann-Whitney U test for continuous variables, Chi-squared or Fisher's exact test for categorical variables, as appropriate.

AR – aortic regurgitation, AS – aortic stenosis, BMI – body mass index, BSA – body surface area, CHD – coronary heart disease, DBP – diastolic blood pressure, DD – diastolic dysfunction, GFR – glomerular filtration rate, HF – heart failure, HR – heart rate, IVSd – thickness of the interventricular septum at end-diastole, LAP – left atrial pressure, LVEDVi – left ventricular end-diastolic volume index, LVESVi – left ventricular end-systolic volume index, LVIDd – left ventricular internal diameter at end-diastole, LVIDs – left ventricular internal diameter at end-systole, LVPWd – thickness of the left ventricular posterior wall at end-diastole, MR – mitral regurgitation, NT-proBNP – N-terminal pro-brain natriuretic peptide, PH – pulmonary hypertension, SBP – systolic blood pressure; other abbreviations as in Supplemental Tables 1 and 2.

**Supplemental Table 4.** Univariable Cox regression models predicting the composite endpoint of HF hospitalization or all-cause death in the ARIC cohort

| | N | HR (95% CI) | P-value |
|---|---|---|---|
| **Demographics and risk factors** | | | |
| Age | 5,450 | 1.106 (1.095–1.117) | <0.001 |
| Male sex | 5,450 | 1.505 (1.351–1.676) | <0.001 |
| Black race | 5,450 | 1.081 (0.938–1.246) | 0.283 |
| Hypertension | 5,450 | 2.006 (1.696–2.372) | <0.001 |
| Diabetes | 5,450 | 1.710 (1.534–1.905) | <0.001 |
| Atrial fibrillation | 5,450 | 2.769 (2.220–3.454) | <0.001 |
| Chronic kidney disease | 5,450 | 1.902 (1.703–2.124) | <0.001 |
| History of HF | 5,450 | 3.267 (2.912–3.666) | <0.001 |
| History of CHD | 5,450 | 2.340 (2.085–2.627) | <0.001 |
| History of non-cardiac PH | 5,450 | 3.761 (2.970–4.763) | <0.001 |
| History of stroke | 5,450 | 2.203 (1.780–2.726) | <0.001 |
| **Laboratory results** | | | |
| Hemoglobin | 5,298 | 0.991 (0.955–1.028) | 0.615 |
| Creatinine | 5,425 | 1.689 (1.587–1.797) | <0.001 |
| GFR | 5,425 | 0.982 (0.979–0.985) | <0.001 |
| $Log_{10}$(NT-proBNP) | 5,287 | 5.410 (4.820–6.071) | <0.001 |
| **Echocardiographic parameters** | | | |
| LVMi | 5,404 | 1.024 (1.022–1.026) | <0.001 |
| LVEDVi | 5,284 | 1.030 (1.026–1.035) | <0.001 |
| LVESVi | 5,284 | 1.057 (1.052–1.063) | <0.001 |
| LVEF | 5,290 | 0.940 (0.934–0.947) | <0.001 |
| LAVi | 5,396 | 1.027 (1.025–1.030) | <0.001 |
| Moderate or greater AS | 5,450 | 3.631 (2.425–5.438) | <0.001 |
| Moderate or greater AR | 5,450 | 2.060 (1.069–3.968) | 0.031 |
| Moderate MR | 5,450 | 2.186 (1.613–2.963) | <0.001 |
| E | 5,450 | 1.013 (1.010–1.016) | <0.001 |
| E-DT | 5,431 | 0.997 (0.996–0.999) | <0.001 |
| A | 5,221 | 1.009 (1.007–1.012) | <0.001 |
| E/A | 5,221 | 1.267 (1.042–1.540) | 0.018 |
| e’ (septal) | 5,450 | 0.819 (0.785–0.853) | <0.001 |
| E/e’ (septal) | 5,450 | 1.087 (1.077–1.096) | <0.001 |
| e’ (lateral) | 5,435 | 0.934 (0.909–0.961) | <0.001 |
| E/e’ (lateral) | 5,435 | 1.079 (1.067–1.092) | <0.001 |
| e’ (average) | 5,435 | 0.867 (0.835–0.900) | <0.001 |
| E/e’ (average) | 5,435 | 1.095 (1.084–1.107) | <0.001 |
| TRV | 3,177 | 3.630 (2.923–4.507) | <0.001 |
| 2016 ASE/EACVI guidelines | 5,450 | | |
| Indeterminate diastolic function | | 1.549 (1.146–2.094) | 0.004 |
| DD with normal LAP | | 2.212 (1.868–2.620) | <0.001 |
| DD with indeterminate LAP | | 3.905 (3.235–4.714) | <0.001 |
| DD with elevated LAP | | 5.624 (4.558–6.938) | <0.001 |

**Supplemental Table 4.** Continued

| | N | HR (95% CI) | P-value |
|---|---|---|---|
| 2025 ASE guidelines | 5,450 | | |
| DD with normal LAP | | 1.593 (1.361–1.865) | <0.001 |
| DD with indeterminate LAP | | 5.067 (3.847–6.674) | <0.001 |
| DD with elevated LAP | | 4.141 (3.423–5.010) | <0.001 |
| DL-predicted probability of DD | 5,450 | 4.632 (4.000–5.364) | <0.001 |
| DL-predicted probability of DD ≥0.50 | 5,450 | 2.744 (2.455–3.067) | <0.001 |

Abbreviations as in Supplemental Tables 1, 2, and 3.

**Supplemental Table 5.** Univariable and multivariable Cox regression models predicting the composite endpoint of HF hospitalization or all-cause death in the ARIC cohort (indeterminate groups excluded)

| | **Univariable models** | | **Multivariable model #1** C-index: 0.731 (0.715–0.747) | | **Multivariable model #2** C-index: 0.723 (0.709–0.737) | | **Multivariable model #3** C-index: 0.739 (0.725–0.753) | |
|---|---|---|---|---|---|---|---|---|
| | HR (95% CI) | P-value | HR (95% CI) | P-value | HR (95% CI) | P-value | HR (95% CI) | P-value |
| Age | 1.106 (1.095–1.117) | <0.001 | 1.087 (1.074–1.101) | <0.001 | 1.084 (1.072–1.096) | <0.001 | 1.075 (1.064–1.087) | <0.001 |
| Male sex | 1.505 (1.351–1.676) | <0.001 | 1.374 (1.207–1.565) | <0.001 | 1.340 (1.195–1.503) | <0.001 | 1.286 (1.150–1.438) | <0.001 |
| Hypertension | 2.006 (1.696–2.372) | <0.001 | 1.176 (0.964–1.435) | 0.110 | 1.144 (0.958–1.366) | 0.136 | 1.078 (0.905–1.285) | 0.400 |
| Diabetes | 1.710 (1.534–1.905) | <0.001 | 1.365 (1.198–1.555) | <0.001 | 1.460 (1.302–1.637) | <0.001 | 1.375 (1.230–1.537) | <0.001 |
| Atrial fibrillation | 2.769 (2.220–3.454) | <0.001 | 1.406 (0.349–5.671) | 0.632 | 1.671 (1.262–2.213) | <0.001 | 1.554 (1.240–1.949) | <0.001 |
| Chronic kidney disease | 1.902 (1.703–2.124) | <0.001 | 1.395 (1.220–1.594) | <0.001 | 1.386 (1.232–1.558) | <0.001 | 1.404 (1.253–1.574) | <0.001 |
| History of HF | 3.267 (2.912–3.666) | <0.001 | 1.918 (1.647–2.233) | <0.001 | 2.106 (1.843–2.407) | <0.001 | 2.037 (1.791–2.317) | <0.001 |
| History of CHD | 2.340 (2.085–2.627) | <0.001 | 1.132 (0.971–1.319) | 0.114 | 1.267 (1.107–1.451) | <0.001 | 1.206 (1.058–1.376) | 0.005 |
| History of non-cardiac PH | 3.761 (2.970–4.763) | <0.001 | 1.836 (1.357–2.485) | <0.001 | 1.155 (0.816–1.634) | 0.416 | 1.690 (1.324–2.157) | <0.001 |
| History of stroke | 2.203 (1.780–2.726) | <0.001 | 1.110 (0.856–1.439) | 0.433 | 1.143 (0.911–1.433) | 0.248 | 1.062 (0.853–1.321) | 0.591 |
| 2016 ASE/EACVI guidelines | | | | | | | | |
| DD with normal LAP | 2.215 (1.871–2.623) | <0.001 | 1.438 (1.198–1.727) | <0.001 | | | | |
| DD with elevated LAP | 5.659 (4.587–6.982) | <0.001 | 2.791 (2.217–3.513) | <0.001 | | | | |
| 2025 ASE guidelines | | | | | | | | |
| DD with normal LAP | 1.594 (1.362–1.866) | <0.001 | | | 1.232 (1.050–1.446) | 0.011 | | |
| DD with elevated LAP | 4.147 (3.427–5.017) | <0.001 | | | 2.216 (1.812–2.711) | <0.001 | | |
| DL-predicted probability of DD | 4.632 (4.000–5.364) | <0.001 | | | | | 2.673 (2.293–3.117) | <0.001 |

Only variables showing a significant association with the outcome in univariable analysis and having no missing values were considered for the multivariable models. Abbreviations as in Supplemental Tables 1, 2, and 3.

**Supplemental Table 6.** Incremental prognostic value of the guideline-based classifications and the DL model over NT-proBNP in the ARIC cohort

| | C-index | ΔC-index | P-value |
|---|---|---|---|
| **NT-proBNP** | 0.703 (0.688–0.718) | | |
| NT-proBNP + **2016 ASE/EACVI guidelines** | 0.715 (0.700–0.730) | +0.012 | <0.001 |
| NT-proBNP + **2025 ASE guidelines** | 0.710 (0.695–0.725) | +0.008 | <0.001 |
| NT-proBNP + **DL-predicted probability of DD** | 0.720 (0.705–0.735) | +0.017 | <0.001 |

Each row corresponds to a Cox regression model predicting the composite endpoint of HF hospitalization or all-cause death, with the variable(s) in the first column included as predictors. The third column shows the difference in the C-index of the given model vs. that of the model containing NT-proBNP only, whereas the P-value in the fourth column refers to the significance of this difference. Abbreviations as in Supplemental Tables 1, 2, and 3.

**Supplemental Table 7.** Prognostic performance of the guidelines and the DL model in the ARIC cohort using time-dependent receiver operating characteristic curves

| | 1st year | 2nd year | 3rd year | 4th year | 5th year | 6th year | 7th year |
|---|---|---|---|---|---|---|---|
| **2016 ASE/EACVI guidelines** | 0.716 (0.677–0.756) | 0.680 (0.652–0.708) | 0.680 (0.657–0.702) | 0.668 (0.649–0.687) | 0.658 (0.641–0.675) | 0.658 (0.641–0.674) | 0.655 (0.634–0.677) |
| **2025 ASE guidelines** | 0.667 (0.625–0.710) | 0.645 (0.617–0.674) | 0.637 (0.614–0.659) | 0.627 (0.608–0.646) | 0.614 (0.597–0.631) | 0.607 (0.591–0.623) | 0.625 (0.606–0.645) |
| **DL model** | 0.787 (0.745–0.829) | 0.741 (0.711–0.771) | 0.729 (0.704–0.753) | 0.719 (0.698–0.740) | 0.702 (0.682–0.721) | 0.691 (0.673–0.709) | 0.710 (0.690–0.731) |
| **DL vs. 2016 guidelines** | p<0.001 | p<0.001 | p<0.001 | p<0.001 | p<0.001 | p<0.001 | p<0.001 |
| **DL vs. 2025 guidelines** | p<0.001 | p<0.001 | p<0.001 | p<0.001 | p<0.001 | p<0.001 | p<0.001 |
| **2025 vs. 2016 guidelines** | p=0.007 | p=0.011 | p<0.001 | p<0.001 | p<0.001 | p<0.001 | p=0.007 |

The composite of HF hospitalization or all-cause death was used as the endpoint in this analysis. Areas under the receiver operating characteristic curve were calculated at the end of each year up to 7 years of follow-up. Abbreviations as in Supplemental Tables 1 and 2.

**Supplemental Table 8.** C-indices of univariable Cox regression models predicting the composite endpoint of HF hospitalization or all-cause death across different subgroups of the ARIC cohort

| | 2016 guidelines | 2025 guidelines | DL model | DL vs. 2016 guidelines | DL vs. 2025 guidelines | 2025 vs. 2016 guidelines |
|---|---|---|---|---|---|---|
| **All participants** | 0.638 (0.624–0.652) | 0.602 (0.588–0.616) | 0.676 (0.660–0.692) | p<0.001 | p<0.001 | p<0.001 |
| **Race** | | | | | | |
| Black (n=1,049) | 0.617 (0.584–0.650) | 0.596 (0.564–0.628) | 0.677 (0.640–0.714) | p<0.001 | p<0.001 | p=0.196 |
| White (n=4,401) | 0.643 (0.627–0.659) | 0.603 (0.588–0.618) | 0.676 (0.659–0.693) | p<0.001 | p<0.001 | p<0.001 |
| **Atrial fibrillation** | | | | | | |
| Yes (n=166) | 0.510 (0.495–0.525) | 0.556 (0.498–0.614) | 0.665 (0.604–0.726) | p<0.001 | p=0.001 | p=0.119 |
| No (n=5,284) | 0.630 (0.615–0.645) | 0.594 (0.580–0.608) | 0.672 (0.656–0.688) | p<0.001 | p<0.001 | p<0.001 |
| **History of HF** | | | | | | |
| Yes (n=908) | 0.600 (0.576–0.624) | 0.612 (0.588–0.636) | 0.689 (0.664–0.714) | p<0.001 | p<0.001 | p=0.292 |
| No (n=4,542) | 0.607 (0.589–0.625) | 0.573 (0.557–0.589) | 0.636 (0.616–0.656) | p=0.004 | p<0.001 | p<0.001 |
| **History of CHD** | | | | | | |
| Yes (n=1,048) | 0.605 (0.581–0.629) | 0.612 (0.588–0.636) | 0.679 (0.653–0.705) | p<0.001 | p<0.001 | p=0.564 |
| No (n=4,402) | 0.621 (0.603–0.639) | 0.586 (0.570–0.602) | 0.655 (0.636–0.674) | p=0.004 | p<0.001 | p<0.001 |
| **Elevated NT-proBNP** | | | | | | |
| Yes (n=1,531) | 0.659 (0.638–0.680) | 0.622 (0.601–0.643) | 0.697 (0.675–0.719) | p<0.001 | p<0.001 | p<0.001 |
| No (n=3,756) | 0.580 (0.561–0.599) | 0.561 (0.543–0.579) | 0.622 (0.600–0.644) | p<0.001 | p<0.001 | p=0.078 |

NT-proBNP levels were considered elevated at ≥125 pg/mL for patients younger than 75 years and ≥450 pg/mL for those aged 75 years or older.

Abbreviations as in Supplemental Tables 1, 2, and 3.

**Supplemental Table 9.** Pairwise comparisons of the event-free survival of patients classified according to the 2016 ASE/EACVI guidelines

| | **Indeterminate diastolic function** | **DD with normal LAP** | **DD with indet. LAP** | **DD with elevated LAP** |
|---|---|---|---|---|
| **Normal diastolic function** | p=0.004 | p<0.001 | p<0.001 | p<0.001 |
| **Indeterminate diastolic function** | | p=0.009 | p<0.001 | p<0.001 |
| **DD with normal LAP** | | | p<0.001 | p<0.001 |
| **DD with indet. LAP** | | | | p<0.001 |

The composite of HF hospitalization or all-cause death was used as the endpoint in this analysis. Cell values are adjusted P-values of pairwise Log-rank tests. P-values were adjusted using the Benjamini-Hochberg method. Abbreviations as in Supplemental Tables 2 and 3.

**Supplemental Table 10.** Pairwise comparisons of the event-free survival of patients classified according to the 2025 ASE guidelines

| | **DD with normal LAP** | **DD with indeterminate LAP** | **DD with elevated LAP** |
|---|---|---|---|
| **Normal diastolic function** | p<0.001 | p<0.001 | p<0.001 |
| **DD with normal LAP** | | p<0.001 | p<0.001 |
| **DD with indeterminate LAP** | | | p=0.150 |

The composite of HF hospitalization or all-cause death was used as the endpoint in this analysis. Cell values are adjusted P-values of pairwise Log-rank tests. P-values were adjusted using the Benjamini-Hochberg method. Abbreviations as in Supplemental Tables 2 and 3.

**Table 11.** Univariable and multivariable Fine-Gray competing risk models predicting HF hospitalization in the ARIC cohort

| | **Univariable models** | | **Multivariable model #1** C-index: 0.770 (0.753–0.787) | | **Multivariable model #2** C-index: 0.765 (0.747–0.783) | | **Multivariable model #3** C-index: 0.788 (0.771–0.805) | |
|---|---|---|---|---|---|---|---|---|
| | sHR (95% CI) | P-value | sHR (95% CI) | P-value | sHR (95% CI) | P-value | sHR (95% CI) | P-value |
| Age | 1.079 (1.064–1.093) | <0.001 | 1.039 (1.024–1.054) | <0.001 | 1.038 (1.023–1.053) | <0.001 | 1.031 (1.016–1.046) | <0.001 |
| Male sex | 1.544 (1.333–1.789) | <0.001 | 1.349 (1.155–1.576) | <0.001 | 1.302 (1.116–1.517) | <0.001 | 1.211 (1.039–1.413) | 0.015 |
| Hypertension | 3.075 (2.339–4.042) | <0.001 | 1.538 (1.160–2.039) | 0.003 | 1.595 (1.202–2.116) | 0.001 | 1.424 (1.070–1.894) | 0.015 |
| Diabetes | 1.963 (1.695–2.273) | <0.001 | 1.431 (1.234–1.660) | <0.001 | 1.424 (1.227–1.653) | <0.001 | 1.341 (1.155–1.557) | <0.001 |
| Atrial fibrillation | 2.071 (2.021–3.608) | <0.001 | 1.040 (0.741–1.458) | 0.822 | 1.168 (0.847–1.611) | 0.344 | 1.170 (0.865–1.583) | 0.309 |
| Chronic kidney disease | 2.052 (1.769–2.380) | <0.001 | 1.381 (1.184–1.612) | <0.001 | 1.428 (1.224–1.666) | <0.001 | 1.424 (1.221–1.660) | <0.001 |
| History of HF | 4.699 (4.055–5.445) | <0.001 | 2.550 (2.161–3.009) | <0.001 | 2.721 (2.308–3.208) | <0.001 | 2.503 (2.125–2.949) | <0.001 |
| History of CHD | 3.005 (2.585–3.494) | <0.001 | 1.362 (1.147–1.618) | <0.001 | 1.453 (1.226–1.722) | <0.001 | 1.323 (1.116–1.569) | 0.001 |
| History of non-cardiac PH | 5.234 (3.973–6.895) | <0.001 | 2.023 (1.496–2.736) | <0.001 | 1.520 (1.097–2.106) | 0.012 | 1.895 (1.404–2.557) | <0.001 |
| History of stroke | 2.424 (1.852–3.174) | <0.001 | 1.077 (0.799–1.451) | 0.625 | 1.036 (0.766–1.402) | 0.818 | 1.003 (0.749–1.345) | 0.982 |
| 2016 ASE/EACVI guidelines | | | | | | | | |
| Indeterminate diastolic function | 1.908 (1.218–2.989) | 0.005 | 1.749 (1.112–2.750) | 0.015 | | | | |
| DD with normal LAP | 2.858 (2.187–3.733) | <0.001 | 1.554 (1.170–2.063) | 0.002 | | | | |
| DD with indeterminate LAP | 5.875 (4.420–7.808) | <0.001 | 2.588 (1.886–3.550) | <0.001 | | | | |
| DD with elevated LAP | 10.136 (7.531–13.643) | <0.001 | 4.213 (3.034–5.850) | <0.001 | | | | |
| 2025 ASE guidelines | | | | | | | | |
| DD with normal LAP | 1.963 (1.539–2.504) | <0.001 | | | 1.585 (1.239–2.028) | <0.001 | | |
| DD with indeterminate LAP | 7.109 (4.886–10.342) | <0.001 | | | 2.267 (1.462–3.517) | <0.001 | | |
| DD with elevated LAP | 7.089 (5.424–9.264) | <0.001 | | | 3.669 (2.761–4.875) | <0.001 | | |
| DL-predicted probability of DD | 9.276 (7.314–11.760) | <0.001 | | | | | 4.931 (3.862–6.296) | <0.001 |

Only variables showing a significant association with the outcome in univariable analysis and having no missing values were considered for the multivariable models. sHR – subdistribution hazard ratio; other abbreviations as in Supplemental Tables 1, 2, and 3.

**Table 12.** Univariable and multivariable Cox regression models predicting all-cause death in the ARIC cohort

| | Univariable models | | Multivariable model #1 C-index: 0.720 (0.703–0.737) | | Multivariable model #2 C-index: 0.717 (0.700–0.734) | | Multivariable model #3 C-index: 0.722 (0.705–0.739) | |
|---|---|---|---|---|---|---|---|---|
| | HR (95% CI) | P-value | HR (95% CI) | P-value | HR (95% CI) | P-value | HR (95% CI) | P-value |
| Age | 1.119 (1.106–1.132) | <0.001 | 1.097 (1.084–1.111) | <0.001 | 1.098 (1.085–1.112) | <0.001 | 1.094 (1.081–1.108) | <0.001 |
| Male sex | 1.544 (1.359–1.754) | <0.001 | 1.402 (1.229–1.600) | <0.001 | 1.389 (1.217–1.584) | <0.001 | 1.351 (1.184–1.541) | <0.001 |
| Hypertension | 1.728 (1.430–2.088) | <0.001 | 1.023 (0.839–1.248) | 0.819 | 1.057 (0.867–1.289) | 0.582 | 1.014 (0.831–1.236) | 0.894 |
| Diabetes | 1.600 (1.409–1.819) | <0.001 | 1.390 (1.218–1.585) | <0.001 | 1.396 (1.224–1.593) | <0.001 | 1.369 (1.200–1.563) | <0.001 |
| Atrial fibrillation | 2.981 (2.333–3.809) | <0.001 | 1.525 (1.143–2.033) | 0.004 | 1.508 (1.140–1.994) | 0.004 | 1.714 (1.333–2.205) | <0.001 |
| Chronic kidney disease | 1.903 (1.671–2.167) | <0.001 | 1.392 (1.217–1.592) | <0.001 | 1.419 (1.241–1.623) | <0.001 | 1.408 (1.231–1.610) | <0.001 |
| History of HF | 2.612 (2.274–3.000) | <0.001 | 1.656 (1.414–1.939) | <0.001 | 1.717 (1.467–2.009) | <0.001 | 1.676 (1.432–1.961) | <0.001 |
| History of CHD | 1.993 (1.736–2.289) | <0.001 | 1.088 (0.928–1.274) | 0.299 | 1.151 (0.984–1.346) | 0.078 | 1.101 (0.941–1.288) | 0.230 |
| History of non-cardiac PH | 3.331 (2.537–4.374) | <0.001 | 1.642 (1.238–2.179) | <0.001 | 1.275 (0.933–1.742) | 0.127 | 1.575 (1.187–2.090) | 0.002 |
| History of stroke | 1.938 (1.495–2.512) | <0.001 | 1.112 (0.853–1.448) | 0.433 | 1.103 (0.847–1.438) | 0.467 | 1.064 (0.817–1.387) | 0.644 |
| 2016 ASE/EACVI guidelines | | | | | | | | |
| Indeterminate diastolic function | 1.469 (1.039–2.078) | 0.030 | 1.245 (0.879–1.765) | 0.218 | | | | |
| DD with normal LAP | 1.951 (1.608–2.368) | <0.001 | 1.335 (1.087–1.640) | 0.006 | | | | |
| DD with indeterminate LAP | 3.350 (2.698–4.159) | <0.001 | 1.715 (1.340–2.195) | <0.001 | | | | |
| DD with elevated LAP | 4.282 (3.349–5.476) | <0.001 | 2.188 (1.681–2.848) | <0.001 | | | | |
| 2025 ASE guidelines | | | | | | | | |
| DD with normal LAP | 1.424 (1.189–1.706) | <0.001 | | | 1.093 (0.910–1.314) | 0.340 | | |
| DD with indeterminate LAP | 5.329 (3.954–7.182) | <0.001 | | | 1.874 (1.296–2.711) | <0.001 | | |
| DD with elevated LAP | 3.067 (2.450–3.839) | <0.001 | | | 1.598 (1.260–2.026) | <0.001 | | |
| DL-predicted probability of DD | 3.346 (2.827–3.960) | <0.001 | | | | | 1.883 (1.575–2.251) | <0.001 |

Only variables showing a significant association with the outcome in univariable analysis and having no missing values were considered for the multivariable models. Abbreviations as in Supplemental Tables 1, 2, and 3.

**Supplemental Table 13.** Clinical and echocardiographic characteristics of patients with elevated and normal invasively measured LVFP in the first hemodynamic validation cohort

| | **Missing** n (%) | **All patients** n=83 | **Elevated LVFP** n=25 | **Normal LVFP** n=58 | **P-value** |
|---|---|---|---|---|---|
| **Demographics, vitals, risk factors** | | | | | |
| Age, years | 0 (0) | 61 (54–66) | 58 (53–64) | 63 (54–67) | 0.306 |
| Male sex | 0 (0) | 56 (67) | 19 (76) | 37 (64) | 0.404 |
| Black race | 0 (0) | 3 (4) | 2 (8) | 1 (2) | 0.214 |
| BMI, kg/m$^2$ | 0 (0) | 30.8 (27.6–34.7) | 33.1 (30.8–37.1) | 29.1 (27.0–34.0) | 0.006 |
| BSA, m$^2$ | 0 (0) | 2.11 (1.94–2.28) | 2.2 (2.09–2.34) | 2.08 (1.92–2.22) | 0.014 |
| SBP, mmHg | 0 (0) | 133 (117–147) | 131 (112–148) | 133 (119–146) | 0.623 |
| DBP, mmHg | 0 (0) | 77 (69–89) | 79 (68–87) | 76.5 (69–90) | 0.732 |
| HR, 1/min | 1 (1) | 73 (65–85) | 83 (73–93) | 68 (62–81) | <0.001 |
| Hypertension | 0 (0) | 82 (99) | 24 (96) | 58 (100) | 0.301 |
| Diabetes | 0 (0) | 42 (51) | 16 (64) | 26 (45) | 0.173 |
| Atrial fibrillation | 0 (0) | 6 (7) | 6 (24) | 0 (0) | <0.001 |
| Chronic kidney disease | 0 (0) | 14 (17) | 7 (28) | 7 (12) | 0.145 |
| History of HF | 0 (0) | 30 (36) | 19 (76) | 11 (19) | <0.001 |
| History of CHD | 0 (0) | 67 (81) | 15 (60) | 52 (90) | 0.005 |
| History of stroke | 0 (0) | 10 (12) | 3 (12) | 7 (12) | 1.000 |
| **Laboratory results** | | | | | |
| Hemoglobin, g/dL | 59 (71) | 11.5 (9.6–12.3) | 11.7 (9.3–12.4) | 10.6 (10.0–11.6) | 0.446 |
| Creatinine, mg/dL | 2 (2) | 1.00 (0.83–1.38) | 1.32 (0.88–1.71) | 0.98 (0.82–1.17) | 0.005 |
| GFR, mL/min/1.73m$^2$ | 2 (2) | 68 (47–87) | 53 (34–86) | 73 (54–88) | 0.031 |
| BNP, pg/mL | 56 (67) | 1,068 (425–2,170) | 1,262 (425–2,206) | 1,027 (462–1,862) | 0.700 |
| **Echocardiographic parameters** | | | | | |
| IVSd, mm | 2 (2) | 11 (11–14) | 11 (10–12) | 12 (11–15) | 0.037 |
| LVPWd, mm | 2 (2) | 10 (9–12) | 11 (9–12) | 9 (8–11) | 0.045 |
| LVIDd, mm | 26 (31) | 46 (42–50) | 52 (47–56) | 45 (42–50) | 0.012 |
| LVIDs, mm | 26 (31) | 33 (29–35) | 34 (33–42) | 32 (29–34) | 0.074 |
| LVMi, g/m$^2$ | 5 (6) | 99.0 (80.8–118.3) | 116.9 (103.5–141.9) | 90.1 (73.7–107.3) | <0.001 |
| LVEDVi, mL/m$^2$ | 2 (2) | 57.3 (47.7–76.0) | 81.2 (62.7–97.3) | 53.1 (44.2–63.9) | <0.001 |
| LVESVi, mL/m$^2$ | 3 (4) | 26.9 (19.6–40.3) | 47.1 (31.2–62.6) | 22.7 (18.6–29.8) | <0.001 |
| LVEF, % | 0 (0) | 54.0 (38.0–59.5) | 32.0 (17.0–47.0) | 56 (51.5–61.8) | <0.001 |
| LAVi, mL/m$^2$ | 3 (4) | 27.9 (20.7–38.7) | 40.7 (35.1–52.9) | 24.55 (20.0–29.6) | <0.001 |
| LARS, % | 18 (22) | 21.4 (11.0–30.5) | 10.7 (7.8–15.0) | 27.0 (18.4–34.1) | <0.001 |
| Moderate or greater AS | 0 (0) | 3 (4) | 1 (4) | 2 (3) | 1.000 |
| Moderate or greater AR | 0 (0) | 1 (1) | 0 (0) | 1 (2) | 1.000 |
| Moderate MR | 0 (0) | 11 (13) | 9 (36) | 2 (3) | <0.001 |
| E, cm/s | 0 (0) | 81 (63–95) | 101 (87–115) | 72 (60–86) | <0.001 |
| E-DT, ms | 1 (1) | 202 (192–232) | 193 (160–208) | 210 (194–235) | 0.009 |
| A, cm/s | 5 (6) | 70 (56–85) | 67 (41–84) | 70 (59–85) | 0.250 |
| E/A | 5 (6) | 1.0 (0.8–1.5) | 1.2 (1.0–2.8) | 1.0 (0.8–1.2) | 0.005 |
| IVRT, ms | 21 (25) | 77 (70–90) | 76 (69–80) | 79 (72–90) | 0.150 |

**Supplemental Table 13.** Continued

| | Missing n (%) | All patients n=83 | Elevated LVFP n=25 | Normal LVFP n=58 | P-value |
|---|---|---|---|---|---|
| e' (septal), cm/s | 0 (0) | 6.0 (4.7–7.3) | 5 (3.3–6.0) | 6.2 (5.0–8.0) | <0.001 |
| E/e' (septal) | 0 (0) | 12.7 (9.5–20.0) | 21.3 (17.0–28.6) | 11.0 (9.1–13.5) | <0.001 |
| e' (lateral), cm/s | 2 (2) | 8.0 (6.0–9.9) | 6 (5.1–9.6) | 8.8 (7.1–9.9) | 0.012 |
| E/e' (lateral) | 2 (2) | 9.1 (6.9–14.6) | 17.3 (12.8–21.7) | 7.9 (6.5–11.5) | <0.001 |
| e' (average), cm/s | 2 (2) | 7.5 (5.5–8.4) | 5.5 (4.7–7.3) | 7.6 (6.2–8.7) | 0.001 |
| E/e' (average) | 2 (2) | 10.1 (8.0–17.8) | 17.9 (15.3–22.8) | 9.5 (7.7–11.9) | <0.001 |
| TRV, m/s | 39 (47) | 2.54 (1.87–2.95) | 2.64 (2.23–3.00) | 2.29 (1.57–2.77) | 0.139 |
| PASP, mmHg | 52 (63) | 24 (13–37) | 37 (15–47) | 21 (12–31) | 0.189 |
| 2016 ASE/EACVI guidelines | 0 (0) | | | | |
| Normal diastolic function | | 1 (1) | 0 (0) | 1 (2) | |
| Indeterminate diastolic function | | 0 (0) | 0 (0) | 0 (0) | |
| DD with normal LAP | | 44 (53) | 4 (16) | 40 (69) | |
| DD with indeterminate LAP | | 16 (19) | 6 (24) | 10 (17) | |
| DD with elevated LAP | | 22 (27) | 15 (60) | 7 (12) | <0.001 |
| 2025 ASE guidelines | 0 (0) | | | | |
| Normal diastolic function | | 34 (41) | 3 (12) | 31 (53) | |
| DD with normal LAP | | 25 (30) | 5 (20) | 20 (34) | |
| DD with indeterminate LAP | | 4 (5) | 3 (12) | 1 (2) | |
| DD with elevated LAP | | 20 (24) | 14 (56) | 6 (10) | <0.001 |
| DL-predicted probability of DD | 0 (0) | 0.95 (0.21–1.00) | 1.00 (1.00–1.00) | 0.80 (0.07–0.99) | <0.001 |
| **Invasive hemodynamic assessment** | | | | | |
| LVFP, mmHg | 0 (0) | 11.1 (7.7–17.5) | 24.0 (18.1–27.0) | 8.7 (5.6–11.1) | <0.001 |

LVFP was measured as PCWP by right heart catheterization or LV pre-atrial contraction pressure by left heart catheterization. Continuous variables are presented as median (interquartile range) and categorical variables as n (%). Comparisons between subgroups with elevated and normal LVFP were performed using unpaired Student's t-test or Mann-Whitney U test for continuous variables, Chi-squared or Fisher's exact test for categorical variables, as appropriate.

PCWP – pulmonary capillary wedge pressure, LV – left ventricular; other abbreviations as in Supplemental Tables 1, 2, and 3.

**Supplemental Table 14.** Clinical and echocardiographic characteristics of patients with elevated and normal invasively measured LVFP in the second hemodynamic validation cohort

| | **Missing** n (%) | **All patients** n=130 | **Elevated LVFP** n=30 | **Normal LVFP** n=100 | **P-value** |
|---|---|---|---|---|---|
| **Demographics, vitals, risk factors** | | | | | |
| Age, years | 0 (0) | 66 (51–75) | 67 (61–72) | 65 (48–75) | 0.241 |
| Male sex | 0 (0) | 70 (54) | 24 (80) | 46 (46) | 0.002 |
| BMI, kg/m$^2$ | 3 (2) | 22.0 (19.4–24.7) | 22.2 (20.1–24.1) | 21.5 (19.3–24.8) | 0.583 |
| BSA, m$^2$ | 3 (2) | 1.58 (1.43–1.73) | 1.65 (1.53–1.76) | 1.55 (1.40–1.73) | 0.055 |
| SBP, mmHg | 0 (0) | 112 (100–126) | 115 (98–126) | 112 (101–127) | 0.916 |
| DBP, mmHg | 0 (0) | 65 (57–76) | 70 (62–75) | 63 (57–76) | 0.250 |
| HR, 1/min | 0 (0) | 70 (62–80) | 72 (67–86) | 70 (61–79) | 0.163 |
| Hypertension | 0 (0) | 49 (38) | 16 (53) | 33 (33) | 0.072 |
| Diabetes | 0 (0) | 30 (23) | 15 (50) | 15 (15) | <0.001 |
| Atrial fibrillation | 0 (0) | 21 (16) | 5 (17) | 16 (16) | 1.000 |
| History of CHD | 0 (0) | 29 (22) | 13 (43) | 16 (16) | 0.004 |
| History of non-cardiac PH | 0 (0) | 41 (32) | 11 (37) | 30 (30) | 0.642 |
| **Laboratory results** | | | | | |
| Hemoglobin, g/dL | 0 (0) | 13.4 (11.7–14.6) | 14.1 (11.0–14.8) | 13.3 (11.8–14.4) | 0.205 |
| BNP, pg/mL | 37 (28) | 168 (45–318) | 315 (161–876) | 113 (33–278) | 0.001 |
| NT-proBNP, pg/mL | 94 (72) | 326 (192–1,282) | 1,971 (321–2,600) | 293 (191–959) | 0.175 |
| **Echocardiographic parameters** | | | | | |
| IVSd, mm | 0 (0) | 10 (9–11) | 11 (9–12) | 10 (9–11) | 0.213 |
| LVPWd, mm | 0 (0) | 10 (9–11) | 10 (8–12) | 10 (9–11) | 0.380 |
| LVIDd, mm | 0 (0) | 47 (41–56) | 56 (47–63) | 46 (41–51) | <0.001 |
| LVIDs, mm | 0 (0) | 31 (26–45) | 48 (34–56) | 29 (25–35) | <0.001 |
| LVMi, g/m$^2$ | 3 (2) | 102.5 (81.5–141.8) | 141.0 (101.9–159.3) | 96.5 (78.4–132.2) | <0.001 |
| LVEDVi, mL/m$^2$ | 103 (79) | 74.2 (56.2–97.5) | 62.7 (39.3–92.7) | 74.2 (57.1–97.5) | 0.389 |
| LVESVi, mL/m$^2$ | 103 (79) | 32.3 (16.3–71.7) | 23.8 (12.7–61.3) | 32.3 (22.0–71.7) | 0.418 |
| LVEF, % | 0 (0) | 61.9 (37.3–69.9) | 31.5 (28.0–49.3) | 64.6 (47.9–71.1) | <0.001 |
| LAVi, mL/m$^2$ | 22 (17) | 35.5 (25.1–48.6) | 39.4 (30.3–57.2) | 33.4 (23.5–46.4) | 0.068 |
| LARS, % | 6 (5) | 17.2 (10.7–27.8) | 9.8 (7.3–14.3) | 21.4 (13.6–32.1) | <0.001 |
| Moderate or greater AS | 40 (31) | 8 (9) | 1 (4) | 7 (11) | 0.430 |
| Moderate or greater AR | 0 (0) | 10 (8) | 1 (3) | 9 (9) | 0.452 |
| Moderate MR | 0 (0) | 21 (16) | 8 (27) | 13 (13) | 0.133 |
| E, cm/s | 0 (0) | 70 (51–90) | 80 (60–104) | 70 (51–85) | 0.103 |
| E-DT, ms | 1 (1) | 192 (150–238) | 171 (146–217) | 195 (159–246) | 0.038 |
| A, cm/s | 22 (17) | 70 (49–84) | 57 (47–83) | 70 (50–86) | 0.448 |
| E/A | 23 (18) | 0.9 (0.6–1.4) | 0.9 (0.6–1.6) | 0.9 (0.6–1.4) | 0.665 |
| IVRT, ms | 75 (58) | 89 (73–114) | 89 (80–108) | 90 (70–113) | 0.880 |
| e’ (septal), cm/s | 0 (0) | 5.5 (3.7–7.3) | 4.3 (3.2–5.2) | 6.0 (4.2–7.6) | <0.001 |
| E/e’ (septal) | 0 (0) | 13.1 (9.7–16.8) | 17.2 (14.0–24.1) | 12.0 (9.0–14.8) | <0.001 |
| e’ (lateral), cm/s | 52 (40) | 8.4 (6.7–11.4) | 6.9 (5.9–8.1) | 9.0 (7.2–11.7) | 0.139 |
| E/e’ (lateral) | 52 (40) | 8.3 (5.9–10.8) | 10.2 (8.6–11.9) | 7.9 (5.8–10.2) | 0.180 |
| e’ (average), cm/s | 52 (40) | 7.3 (5.6–9.2) | 5.5 (5.0–6.2) | 7.8 (6.4–9.6) | 0.092 |
| E/e’ (average) | 52 (40) | 9.2 (7.2–11.9) | 11.1 (9.5–14.3) | 8.9 (7.1–11.3) | 0.068 |

**Supplemental Table 14.** Continued

| | Missing n (%) | All patients n=130 | Elevated LVFP n=30 | Normal LVFP n=100 | P-value |
|---|---|---|---|---|---|
| TRV, m/s | 5 (4) | 2.69 (2.24–3.20) | 2.84 (2.42–3.10) | 2.60 (2.20–3.20) | 0.407 |
| PASP, mmHg | 63 (48) | 37 (26–49) | 44 (37–59) | 37 (26–48) | 0.259 |
| 2016 ASE/EACVI guidelines | 0 (0) | | | | |
| Normal diastolic function | | 9 (7) | 0 (0) | 9 (9) | |
| Indeterminate diastolic function | | 6 (5) | 0 (0) | 6 (6) | |
| DD with normal LAP | | 52 (40) | 11 (37) | 41 (41) | |
| DD with indeterminate LAP | | 33 (25) | 9 (30) | 24 (24) | |
| DD with elevated LAP | | 30 (23) | 10 (33) | 20 (20) | 0.172 |
| 2025 ASE guidelines | 0 (0) | | | | |
| Normal diastolic function | | 9 (7) | 1 (3) | 8 (8) | |
| DD with normal LAP | | 59 (45) | 6 (20) | 53 (53) | |
| DD with indeterminate LAP | | 12 (9) | 4 (13) | 8 (8) | |
| DD with elevated LAP | | 50 (38) | 19 (63) | 31 | 0.003 |
| DL-predicted probability of DD | 0 (0) | 0.98 (0.60–0.99) | 0.99 (0.98–0.99) | 0.93 (0.24–0.99) | <0.001 |
| **Invasive hemodynamic assessment** | | | | | |
| PCWP, mmHg | 0 (0) | 10.0 (7.0–14.8) | 20.0 (17.0–25.8) | 8.0 (5.8–11.0) | <0.001 |

Continuous variables are presented as median (interquartile range) and categorical variables as n (%). Comparisons between subgroups with elevated and normal LVFP were performed using unpaired Student's t-test or Mann-Whitney U test for continuous variables, Chi-squared or Fisher's exact test for categorical variables, as appropriate.

Abbreviations as in Supplemental Tables 1, 2, and 3.

**Supplemental Figure 1.** Prognostic value of the guidelines and the DL model in multivariable Cox regression analysis for predicting the composite endpoint

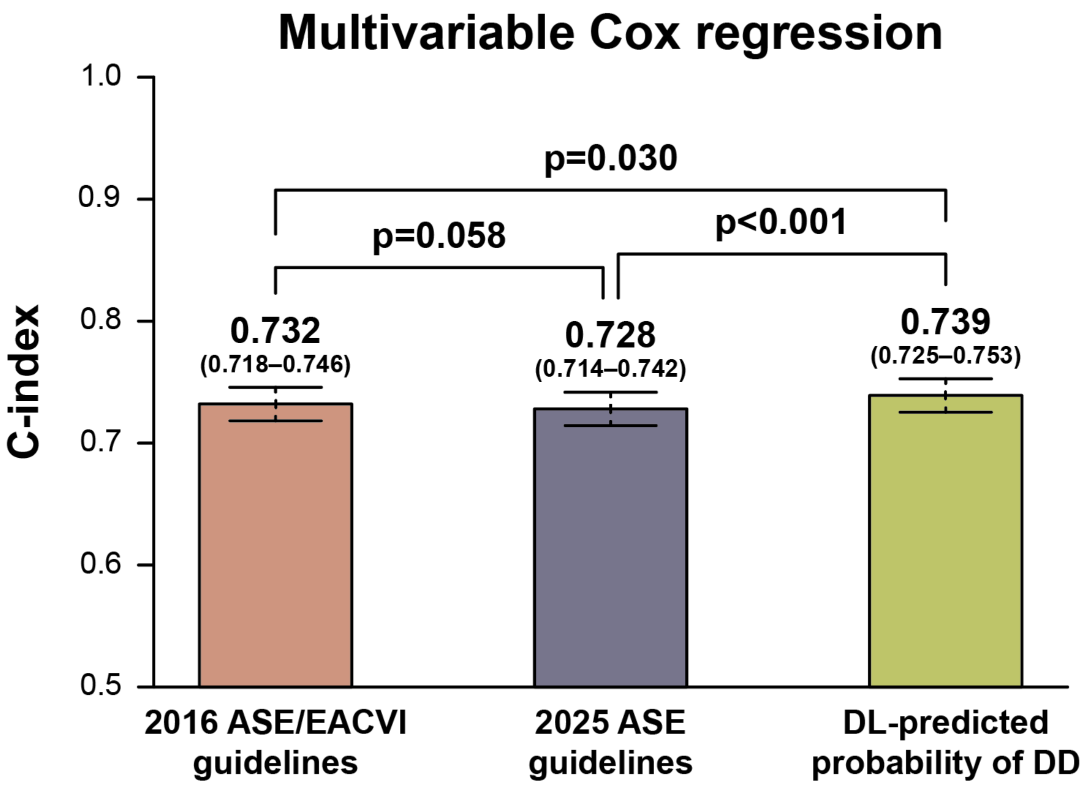


C-indices were calculated from multivariable Cox regression models that included age, sex, hypertension, diabetes, atrial fibrillation, chronic kidney disease, history of heart failure, history of coronary heart disease, history of non-cardiac pulmonary hypertension, and history of stroke, along with either the 2016 guideline-based classification, the 2025 guideline-based classification, or the DL-predicted probability of DD.

ASE – American Society of Echocardiography, EACVI – European Association of Cardiovascular Imaging, DD – diastolic dysfunction, DL – deep learning

**Supplemental Figure 2.** Time-dependent ROC curves of the guidelines and the DL model for predicting the composite endpoint of HF hospitalization or all-cause death

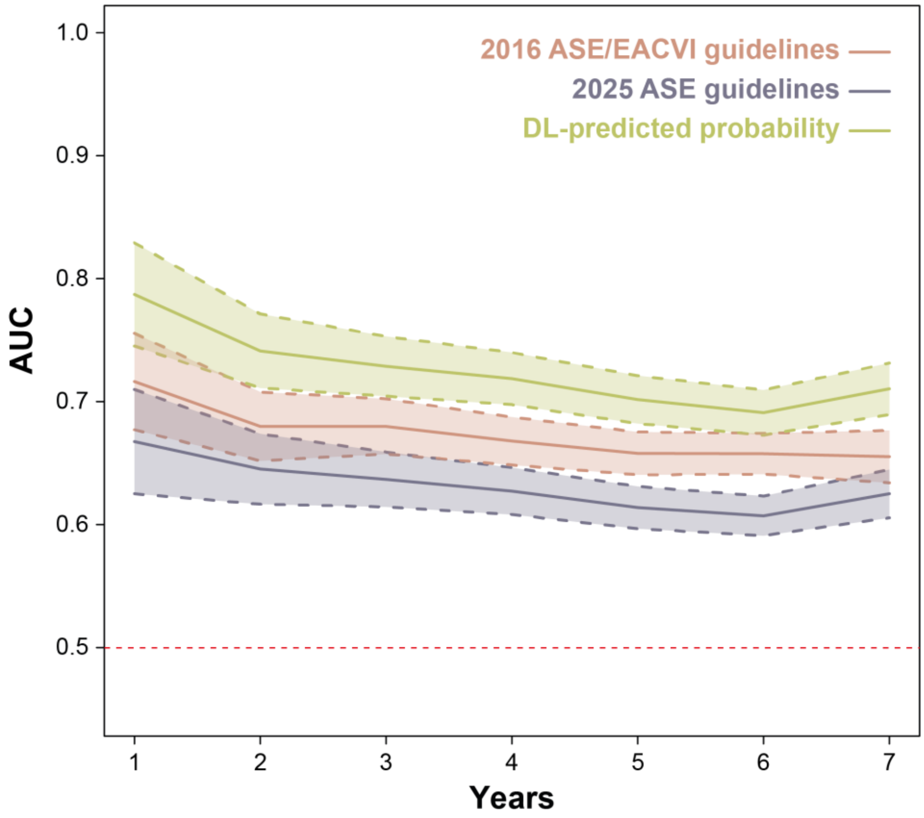


Cumulative time-dependent ROC curves were estimated using inverse probability of censoring weighting and are plotted with 95% confidence bands.

AUC – area under the receiver operating characteristic curve, ROC – receiver operating characteristic; other abbreviations as in Supplemental Figure 1.

**Supplemental Figure 3.** Prognostic value of the guidelines and the DL model in univariable analyses for predicting HF hospitalization and all-cause death separately

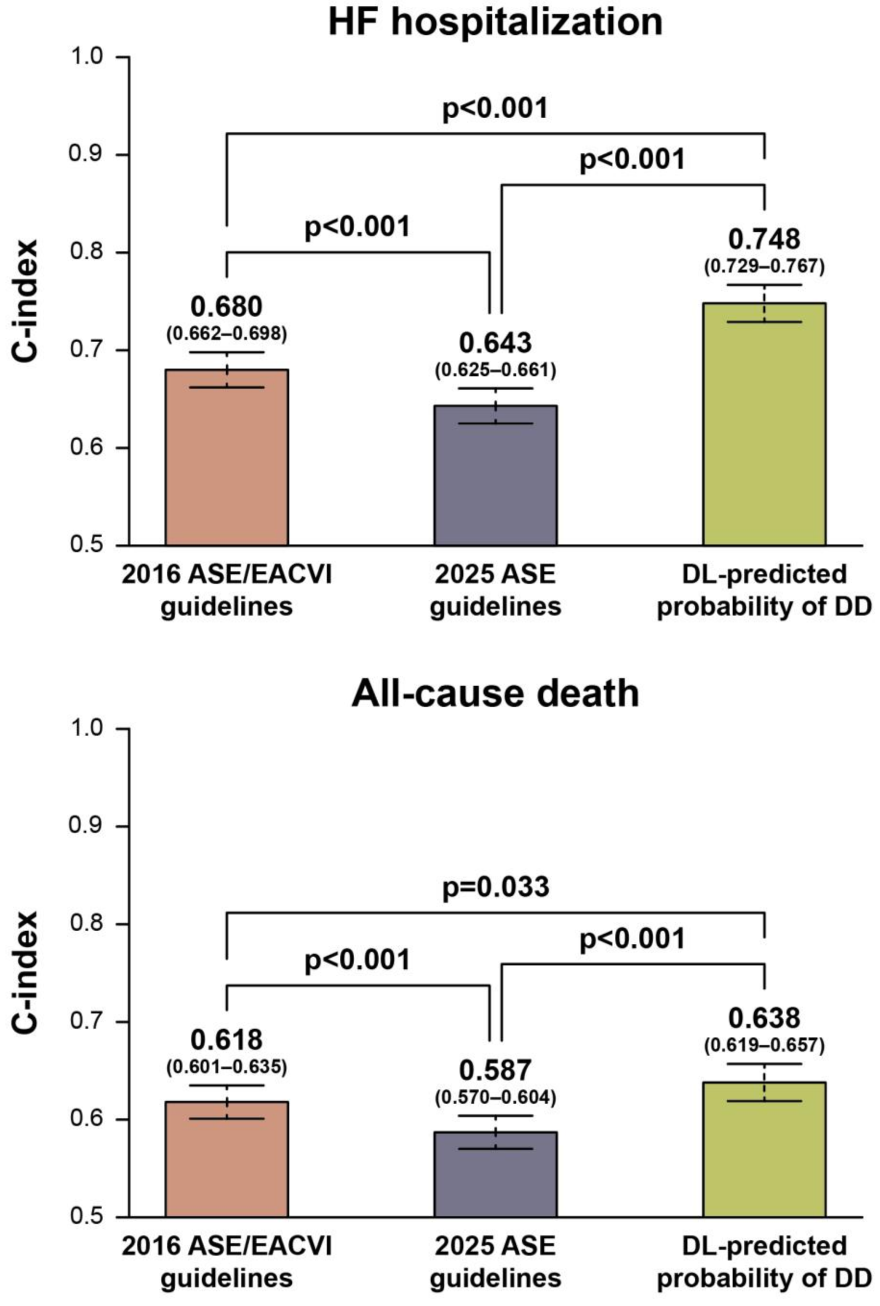


For HF hospitalization, C-indices were calculated from univariable Fine-Gray competing risk regression models (with all-cause death treated as the competing event) that included either the 2016 or 2025 guideline-based classification or the DL-predicted probability of DD as the sole predictor. For all-cause death, C-indices were calculated from univariable Cox regression models using the same predictor structure.

HF – heart failure; other abbreviations as in Supplemental Figure 1.

**Supplemental Figure 4.** ROC curves showing the performance of the guidelines and the DL model in detecting elevated LVFP after excluding patients with atrial fibrillation

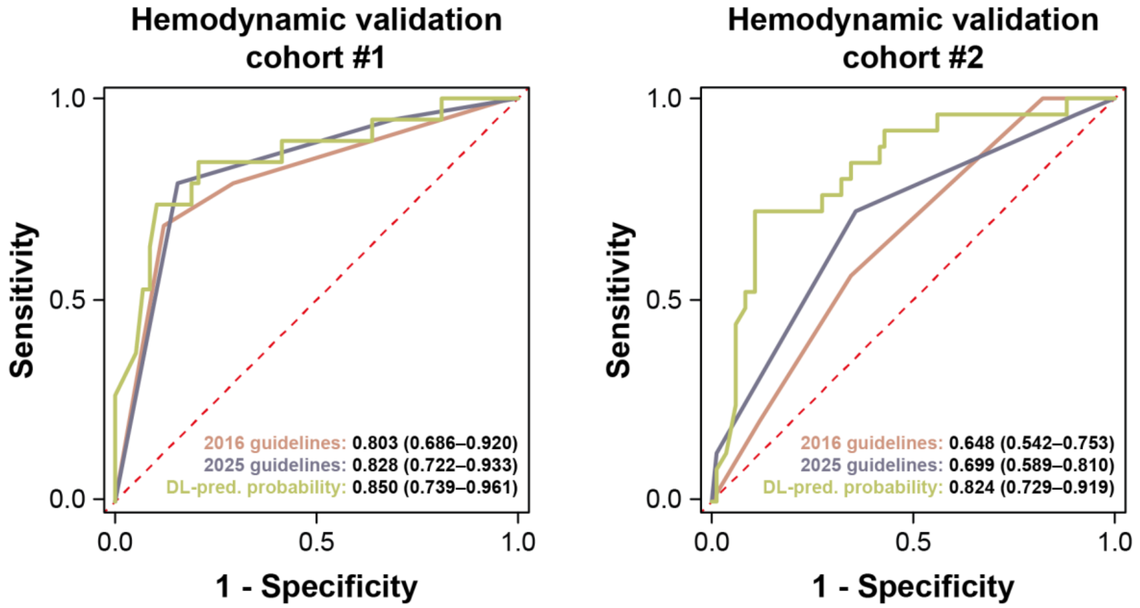


In this sensitivity analysis, 6 of 83 and 21 of 130 patients with atrial fibrillation were excluded from the first and second hemodynamic validation cohorts, respectively. AUCs are reported with 95% CIs.

LVFP – left ventricular filling pressure; other abbreviations as in Supplemental Figures 1 and 2.

**Supplemental Figure 5.** ROC curves showing the performance of the guidelines and the DL model in detecting elevated LVFP after excluding patients with both IVRT and LARS missing

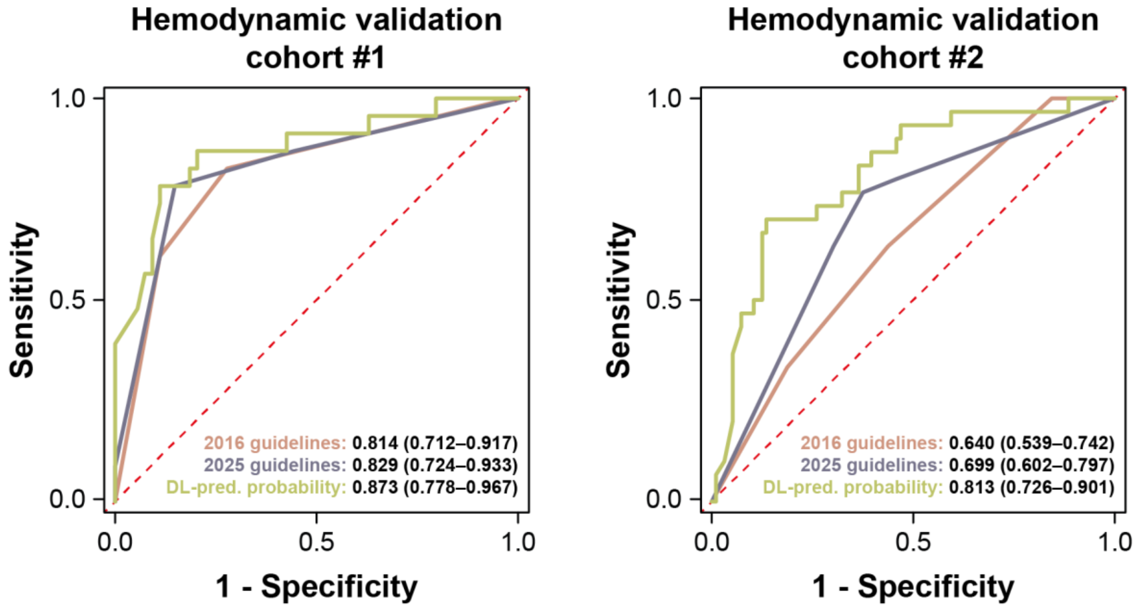


In this sensitivity analysis, 6 of 83 and 4 of 130 patients with both IVRT and LARS missing were excluded from the first and second hemodynamic validation cohorts, respectively. AUCs are reported with 95% CIs.

IVRT – isovolumic relaxation time, LARS – left atrial reservoir strain; other abbreviations as in Supplemental Figures 1, 2, and 4.
